# Occupational endotoxin exposure, DNA methylation changes, and immune-cell remodeling: an epigenome-wide association study in two cohorts

**DOI:** 10.64898/2026.09.29.26364351

**Authors:** Hantao Wang, Mohammad L. Rahman, Charles E. Breeze, Gabriel Goodney, Jungeun Lim, Wei Hu, Batel Blechter, Jianxin Shi, Fengying Zhang, Qiuyin Cai, Gong Yang, Jirong Long, Li Su, Yu-Tang Gao, Xiao-Ou Shu, Nathaniel Rothman, Wei Zheng, Qing Lan, David C. Christiani, Jason Y.Y. Wong

## Abstract

**Background:** Millions of textile and agricultural workers inhale organic dust containing endotoxin, an exposure associated with impaired respiratory function. However, the underlying biological pathways remain unclear. We conducted a two-cohort epigenome-wide association study (EWAS) to identify airborne endotoxin-associated DNA methylation (DNAm) alterations and pathways.

**Methods:** In the discovery phase, we compared blood DNAm in 281 endotoxin-exposed cotton workers and 253 unexposed silk-workers from the Shanghai Textile Worker Study (STWS) measured using Illumina MethylationEPICv2 arrays. STWS endotoxin assessment combined detailed work histories and onsite measurements. For targeted parallel validation, we tested STWS signals in 686 incident lung cancer cases and 683 matched controls from the Shanghai Women’s Health Study (SWHS), using occupation-derived low, medium, or high exposure-intensity ranks based on published evidence. To gain biological insight, we analyzed pathway enrichment, immune-cell composition, DNAm inflammation risk scores (IRSs), and respiratory phenotype associations in STWS.

**Results:** In STWS, we identified 36 epigenome-wide significant DNAm sites (P<5×10⁻⁸), of which, 35 were hypomethylated. Among 30 sites testable in SWHS, 18 had concordant effect directions, and four were nominally significant: cg11869499 (*POLG*; P=0.041), cg16290931 (*TSKS*; P=0.044), cg05019530 (*EPIC1*; P=0.042), and cg26033526 (*PSMB9/TAP1*; P=0.013). Enrichment implicated immune and signal-transduction pathways, including IgSF cell-adhesion-molecule signaling (FDR=8.2×10⁻⁶). Neutrophils were higher in cotton workers than in silk workers (63.5% vs 58.9%, p=7.52×10⁻^10^). Cumulative endotoxin was associated with higher CRP-based IRSs, which showed the most consistent associations with lower lung function and airflow obstruction.

**Conclusions:** Our EWAS findings suggest that occupational endotoxin may operate partly through immune-cell composition and inflammatory pathways.

## Introduction

Endotoxin, or lipopolysaccharide (LPS), is a biologically active component of the outer membrane of Gram-negative bacteria released during bacterial growth and lysis. Endotoxin is common in occupational environments, with particularly high concentrations in the textile and agricultural industries. Workers who handle raw plant materials, livestock, or organic dusts are exposed mainly through inhalation. Among cotton textile workers and other exposed groups, endotoxin has been associated with acute and chronic respiratory effects (1–9).

Endotoxin affects the respiratory system largely through immune activation and inflammation. The CD14/MD-2/Toll-like receptor 4 (TLR4) complex recognizes LPS and activates MyD88- and TRIF-dependent signaling, leading to the production of proinflammatory cytokines (10–12). Previous analyses in cotton textile workers suggested that adaptive immunologic proteins may contribute to endotoxin-related lung function decline (9). However, the molecular pathways linking cumulative exposure to immune remodeling and subsequent respiratory impairment remain unclear.

Blood DNA methylation (DNAm) patterns can reflect environmental exposures (13, 14) and cellular states relevant to chronic lung disease (15). Many cytosine-phosphate-guanine (CpG) sites have immune-cell-specific methylation profiles, allowing immune-cell composition and exposure-related immune remodeling to be characterized from whole-blood DNAm (16).

Furthermore, DNAm-derived inflammation risk scores (IRSs) can capture broader systemic inflammatory processes (17–20). Although experimental studies have shown that LPS can alter epigenetic regulation (21), little is known about the blood methylome response to long-term endotoxin exposure in humans. Examining this response may help identify specific biological pathways linking cumulative exposure with respiratory impairment.

To investigate molecular responses to occupational endotoxin exposure, we analyzed blood methylome data from two independent prospective cohorts. In the discovery phase, we conducted an untargeted epigenome-wide association study (EWAS) in the Shanghai Textile Worker Study (STWS) to identify differentially methylated positions (DMPs) associated with occupational exposure to endotoxin. Subsequently, we conducted targeted parallel validation of the identified signals (referred to as “validation” for brevity) in the population-based Shanghai Women’s Health Study (SWHS) (22–25) using occupation-derived ordinal exposure-intensity rank. We also examined pathway enrichment, immune-cell composition, and DNAm-derived IRSs to characterize endotoxin-related immune remodeling. Finally, we explored associations between endotoxin-related DNAm biomarkers and respiratory phenotypes in the STWS.

## Methods

### Study populations

#### Discovery EWAS Phase

The study design, questionnaires, endotoxin exposure assessment and outcome assessment in STWS have been described previously (1–9). Briefly, the STWS is a longitudinal occupational cohort that prospectively followed 447 endotoxin-exposed cotton workers and 472 unexposed control silk workers in Shanghai, China from 1981 to 2016, with follow-up surveys approximately every 5 years. Blood samples collected in 1996 were available for 284 cotton and 255 silk workers. The Institutional Review Boards (IRBs) of the Harvard T.H. Chan School of Public Health and Putuo District Hospital approved the STWS.

#### Targeted Parallel Validation

The SWHS is a prospective cohort that enrolled 74,941 women aged 40–70 years from Shanghai, China between 1996 and 2000 (22, 23). Participants completed in-person interviews at enrollment and follow-up that collected demographic, lifestyle, and occupational information (24). We used a nested case-control sample of incident lung cancer cases and individually matched controls (25). Controls were alive and at risk on each case’s index date and were matched by date of birth (±2 years) and date of blood collection (±3 months). All participants in this subset were never-smokers and had provided pre-diagnostic blood samples at enrollment. The SWHS was approved by IRBs at the Vanderbilt University Medical Center and Shanghai Cancer Institute.

### Endotoxin Exposure Assessment

#### Discovery EWAS Phase

The primary STWS exposure contrasted cotton workers with silk-worker controls. Full-shift area samples of cotton dust were collected onsite at every survey prior to 2001 using vertical elutriators. Endotoxin concentrations were measured using the Limulus Amebocyte Lysate assay. Previous validation showed that area measurements correlated closely with personal measurements. Samples from silk mills contained nearly undetectable endotoxin concentrations (<0.001 EU/m^3^) (26, 27).

Cumulative endotoxin concentration (EU/m³-year) and cotton dust concentration (mg/m³-year) were estimated using onsite samplings, job titles, and work history. Average 8-hour endotoxin and dust concentrations were calculated from cumulative exposure and years worked. Years since cessation of endotoxin exposure were derived from work history and were set to zero for silk workers. To examine the exposure-response pattern, we defined a four-level ordinal exposure variable with silk workers as the reference group and cotton workers classified by average 8-hour endotoxin concentration as <90, 90 to <1,700, or ≥1,700 EU/m^3^.

#### Targeted Parallel Validation

Each participant’s occupation was collected in questionnaires and coded according to the Chinese National Standard Occupation and Industry Codes Manual. We used each occupation code to assign an occupation-derived exposure-intensity rank. This rank represented relative occupational exposure potential rather than individually measured endotoxin concentrations. Occupations were first grouped into broad occupational categories and assigned low, medium, or high ordinal exposure-intensity rank based on published exposure literature (Detailed in Supplemental Table 1 and 2). An occupation’s rank was raised above its broad-group assignment when its usual tasks were expected to involve more frequent or direct contact with the principal carriers of endotoxin, and lowered when such contact was expected to be less (28).

### Epigenome-wide DNA methylation profiling

#### Discovery EWAS Phase

DNAm was profiled in bisulfite-converted genomic DNA extracted from banked peripheral blood using the Illumina Infinium MethylationEPIC v2.0 BeadChip (EPIC v2; 930K) (29). DNAm data pre-processing and quality control (QC) were performed using SeSAMe (30), including restriction to standard CpG probes, Infinium I channel inference, nonlinear dye-bias correction, pOOBAH-based detection assessment, noob background correction, and Infinium I/II design-bias correction. We then excluded CpG probes with ≥50% missing values and samples with >10% missing values, applied the array-specific EPIC v2 general mask, and removed sex-chromosome probes and beta-distribution outliers. We imputed remaining missing values using k-nearest neighbors (k=10). We then used ComBat (31) to adjust for batch defined by the interaction of Sample Well and Sentrix ID while preserving occupation group, sex, and smoking status. After mapping EPIC v2 probes to EPIC-compatible probes, we estimated immune-cell fractions with EpiDISH (16). Before the EWAS, we removed samples with duplicated participant ID.

#### Targeted Parallel Validation

The National Cancer Institute Cancer Genomics Research Laboratory profiled DNAm in peripheral-blood DNA using the Infinium MethylationEPIC BeadChip (EPIC v1; 850K) (32). QC has been detailed previously (25). ComBat was used for batch-effect correction (31), with Illumina slide as the batch variable. We tested only signals identified in the STWS discovery phase. Probe identifiers were harmonized between EPIC v1 and v2 arrays using the mLiftover function in SeSAMe (33).

### Respiratory outcome assessment

In the STWS, forced expiratory volume in 1 second (FEV-1) and forced vital capacity (FVC) were measured approximately every 5 years following American Thoracic Society spirometry standards (34). Percent-predicted FEV-1 and FVC and the lower limit of normal (LLN) for FEV-1/FVC were calculated using Global Lung Function Initiative (GLI) race-neutral reference equations (35, 36). Annual change in FEV-1 and FVC were calculated between consecutive surveys. Respiratory symptoms were collected using self-reported physician-diagnosis questionnaires at each survey.

### Statistical Analysis

Figure 1 summarizes the study design and analysis workflow.

**Figure 1.**
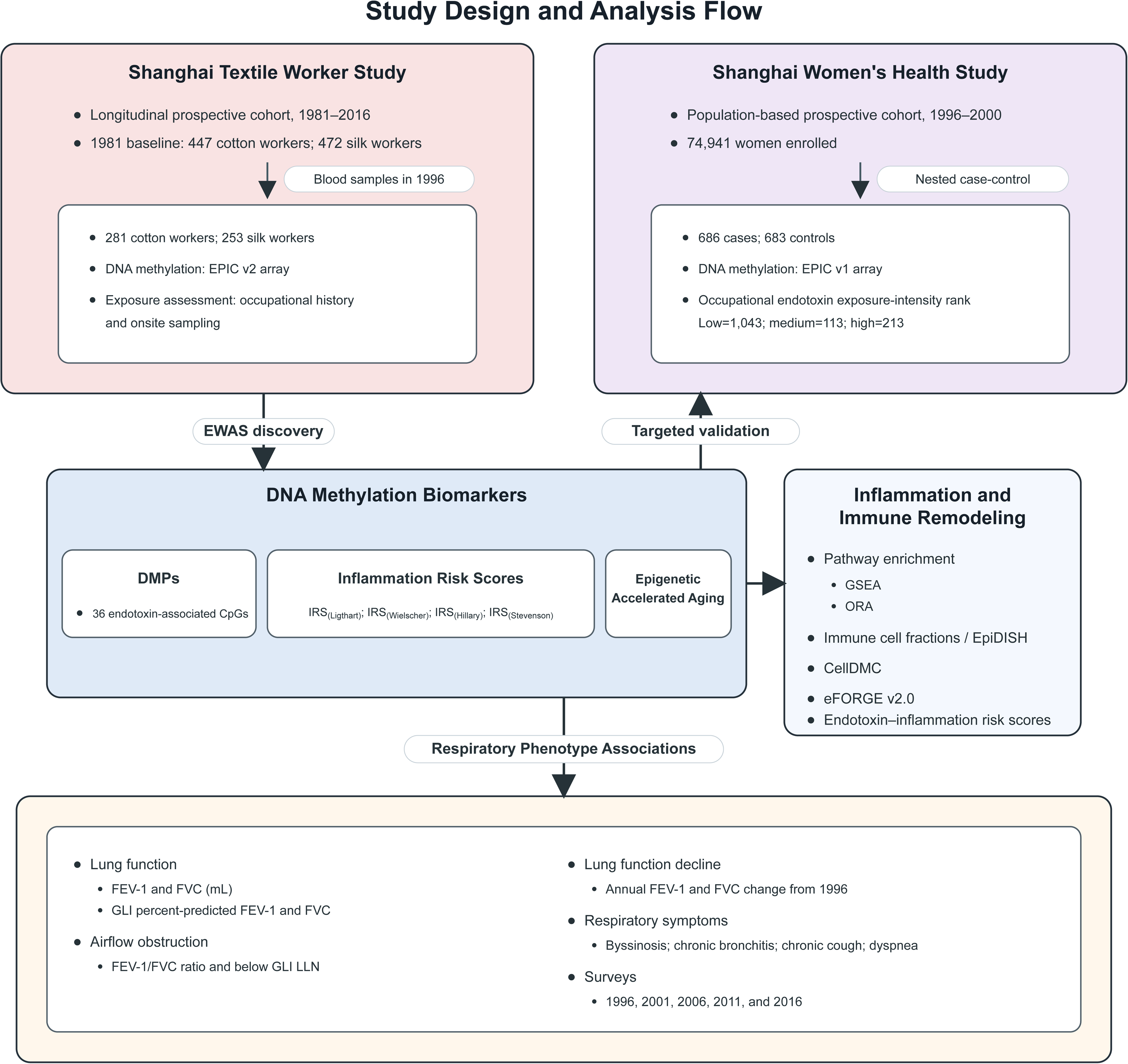
Study design and statistical analysis. This study aimed to identify DNA methylation changes associated with occupational exposure to endotoxin. STWS participants with blood samples collected in 1996 formed the EWAS discovery sample; a nested case-control sample from SWHS provided targeted validation. Endotoxin-associated DNA methylation biomarkers were evaluated for epigenetic accelerated aging, inflammation risk, immune remodeling, and respiratory phenotype associations. **Abbreviations:** CellDMC=cell-type-specific differential methylation; CpG=cytosine-phosphate-guanine; DMP=differentially methylated position; eFORGE v2.0=experimentally derived Functional element Overlap analysis of ReGions from EWAS, version 2.0; EPIC=Infinium MethylationEPIC array; EpiDISH=Epigenetic Dissection of Intra-Sample Heterogeneity; EWAS=epigenome-wide association study; FEV-1=forced expiratory volume in 1 second; FVC=forced vital capacity; GLI=Global Lung Function Initiative; GSEA=Gene Set Enrichment Analysis; IRS=inflammation risk score; LLN=lower limit of normal; ORA=Over-Representation Analysis; STWS=Shanghai Textile Worker Study; SWHS=Shanghai Women’s Health Study.

### Discovery Phase in STWS

In the discovery EWAS, we used linear mixed models to regress DNAm beta values on occupational group, comparing endotoxin-exposed cotton workers with silk-worker controls. Models adjusted for age, sex, body mass index (BMI), smoking pack-years, years since exposure cessation, and EpiDISH-estimated immune-cell fractions. Nested random intercepts for sample plate and Sentrix ID accounted for residual technical effects. We used robust sandwich variance estimators to address correlated observations and heteroskedasticity.

Epigenome-wide significance was defined as p<5×10^-8^. DMPs were excluded if P>0.05 or inconsistent effect directions in sensitivity analyses using simple linear regression. To assess possible nonlinearity, we repeated the analysis using a four-level ordinal endotoxin-exposure variable. Additional sensitivity analyses used cumulative endotoxin and cumulative dust as alternative exposure metrics. We classified DMPs as potentially reversible when years since exposure cessation was associated with methylation at P<0.05 in the direction opposite to the endotoxin effect.

We also examined published DNAm-derived IRSs(17–20), and epigenetic age acceleration (EAA) estimated with dnaMethyAge (37). The construction of IRS_(Ligthart)_, IRS_(Wielscher)_, IRS_(Hillary)_, and IRS_(Stevenson)_ has been previously reported (25). We regressed each standardized IRS and EAA on log-transformed cumulative endotoxin in separate models adjusted for age, sex, BMI, smoking pack-years, years since exposure cessation, and EpiDISH-estimated immune-cell fractions.

### Targeted Parallel Validation in SWHS

DMPs identified in STWS were tested in SWHS using occupation-derived ordinal exposure-intensity ranks. The main validation model compared high vs. low and medium ordinal exposure-intensity rank, with age, BMI, education, case-control status, immune cell fractions, sample well, and a case-control pair random intercept. Sensitivity analyses included inverse probability weighting (IPW) for case-control status and alternative exposure metrics. Associations of ordinal exposure-intensity rank with EAA and IRS were evaluated using linear mixed models adjusted for the same covariates.

### Pathway enrichment and immune-cell remodeling

Pathway enrichment was evaluated using gene set enrichment analysis (GSEA) of ranked gene-level significance (38) against Gene Ontology (GO) terms, Kyoto Encyclopedia of Genes and Genomes (KEGG) pathways, and Reactome pathways. We considered gene sets with a false discovery rate (FDR) <0.05 significant and collapsed related sets into representative pathways. We then summarized leading-edge genes that contributed to multiple enriched pathways. In parallel, we used over-representation analysis (ORA) to identify pathways represented among genes mapped to the endotoxin-associated DMPs (39).

To characterize immune-cell remodeling associated with occupational endotoxin exposure, we examined immune-cell composition, cell-type-specific methylation, and regulatory enrichment. Immune-cell fractions between cotton and silk workers were compared using t-tests. We used CellDMC to identify cell-type-specific differential methylation associated with occupations (40) and eFORGE v2.0 (41) to test whether the endotoxin-associated DMPs overlapped tissue- or cell-type-specific regulatory annotations.

### Targeted DNAm biomarker and respiratory phenotype analyses

To evaluate whether endotoxin-related DNAm biomarkers were associated with respiratory phenotypes, we evaluated associations of endotoxin-related DMPs and IRSs with selected concurrent and prospective lung-function and airflow phenotypes. Models adjusted for log cumulative endotoxin, baseline age, sex, height, years since exposure cessation, smoking pack-years, BMI, immune-cell fractions, and sample plate. Age, sex, and height were omitted for percent predicted FEV-1 and FVC. DMP associations were expressed per 1-SD higher methylation beta value. To assess longitudinal lung function trajectories, we used linear mixed models to estimate associations of 1996 IRSs with FEV-1 and FVC from 1996 through 2016, measured at approximately 5-year intervals, with IPW accounting for loss to follow-up and random intercepts for individual variability. Additional IRS-associated lung function change over follow-up was assessed separately using interactions of IRS and time (*P*trend).

## Results

### Participant characteristics

#### Discovery EWAS Phase

After sample-level QC and removal of duplicate records, the final STWS analytic sample included 281 cotton workers and 253 silk workers (Supplemental Figure 1). Cotton and silk workers were quantitatively similar in age, sex distribution, smoking patterns, and BMI (Table 1). Cotton workers had more respiratory symptoms, slightly lower percent-predicted FEV-1, and greater annual FEV-1 decline.

**Table 1.** Participant Characteristics of Shanghai Textile Worker Study (STWS) in 1996.

| Variable | Cotton | Silk |
| --- | --- | --- |
| **Sex** |  |  |
| Female, % | 165 (58.7%) | 151 (59.7%) |
| Male, % | 116 (41.3%) | 102 (40.3%) |
| **Age, year** |  |  |
| Mean (SD) | 51.6 (10.2) | 51.6 (10.1) |
| **Years worked, year** |  |  |
| Mean (SD) | 26.0 (8.0) | 27.4 (9.2) |
| **Cumulative endotoxin, 1000 EU/m<sup>3</sup>-year** |  |  |
| Mean (SD) | 50.0 (45.0) | / |
| **Cumulative dust, mg/m<sup>3</sup>-year** |  |  |
| Mean (SD) | 18.6 (12.7) | / |
| **Smoking status** |  |  |
| Never, % | 183 (65.1%) | 172 (68.0%) |
| Former, % | 84 (29.9%) | 67 (26.5%) |
| Current, % | 14 (5.0%) | 14 (5.5%) |
| **Smoking pack-years, pack-year** |  |  |
| Mean (SD) | 5.4 (10.5) | 4.9 (10.2) |
| **Body mass index, kg/m<sup>2</sup>** |  |  |
| Mean (SD) | 23.7 (3.2) | 23.6 (2.9) |
| **Symptoms** |  |  |
| Byssinosis, % | 37 (13.2%) | 0 (0.0%) |
| Chronic bronchitis, % | 12 (4.3%) | 4 (1.6%) |
| Chronic cough, % | 9 (3.2%) | 4 (1.6%) |
| Dyspnea, % | 31 (11.0%) | 18 (7.1%) |
| **FEV-1, ml** |  |  |
| Mean (SD) | 2438 (675) | 2413 (593) |
| **% predicted FEV-1, %** |  |  |
| Mean (SD) | 86.2 (14.4) | 87.3 (13.3) |
| **FVC, ml** |  |  |
| Mean (SD) | 3191 (780) | 3157 (725) |
| **% predicted FVC, %** |  |  |
| Mean (SD) | 91.6 (12.1) | 92.6 (10.8) |
| **Annual change in FEV-1 between 1992 and 1996, ml/year** |  |  |
| Mean (SD) | -41.5 (39.6) | -34.4 (34.1) |
| **Annual change in FVC between 1992 and 1996, ml/year** |  |  |
| Mean (SD) | -28.7 (57.6) | -15.1 (41.7) |
| **FEV-1/FVC ratio** |  |  |
| Mean (SD) | 0.762 (0.089) | 0.765 (0.087) |

Abbreviations: EU=endotoxin unit; FEV-1=forced expiratory volume in 1 second; FVC=forced vital capacity; SD=standard deviation; STWS=Shanghai Textile Worker Study. / indicates not available or not applicable.

#### Targeted Parallel Validation

The SWHS subset included 1,369 participants, with 686 incident lung cancer cases and 683 matched controls (Supplemental Table 3). Overall, 214 participants (15.6%) worked in textile occupations. The ordinal exposure-intensity rank was high for 213 participants (15.6%), medium for 113 (8.3%), and low for 1,043 (76.2%). Exposure-intensity rank did not differ by case-control status (chi-square P=0.54)

### DNAm biomarkers associated with occupational endotoxin exposure

#### Discovery in STWS

We identified 36 DMPs associated with occupational endotoxin exposure when comparing cotton and silk workers (Table 2). The three strongest signals were cg22740441 (*PPP3CA*; P=4.76 × 10⁻¹¹), cg11248361 (*AQP4-AS1*; P=1.75 × 10⁻¹⁰), and cg08009902 (*MTA3*; P=6.05 × 10⁻¹⁰). Thirty-five of the 36 DMPs were hypomethylated. Supplemental Figure 2 shows their genomic locations.

**Table 2.**
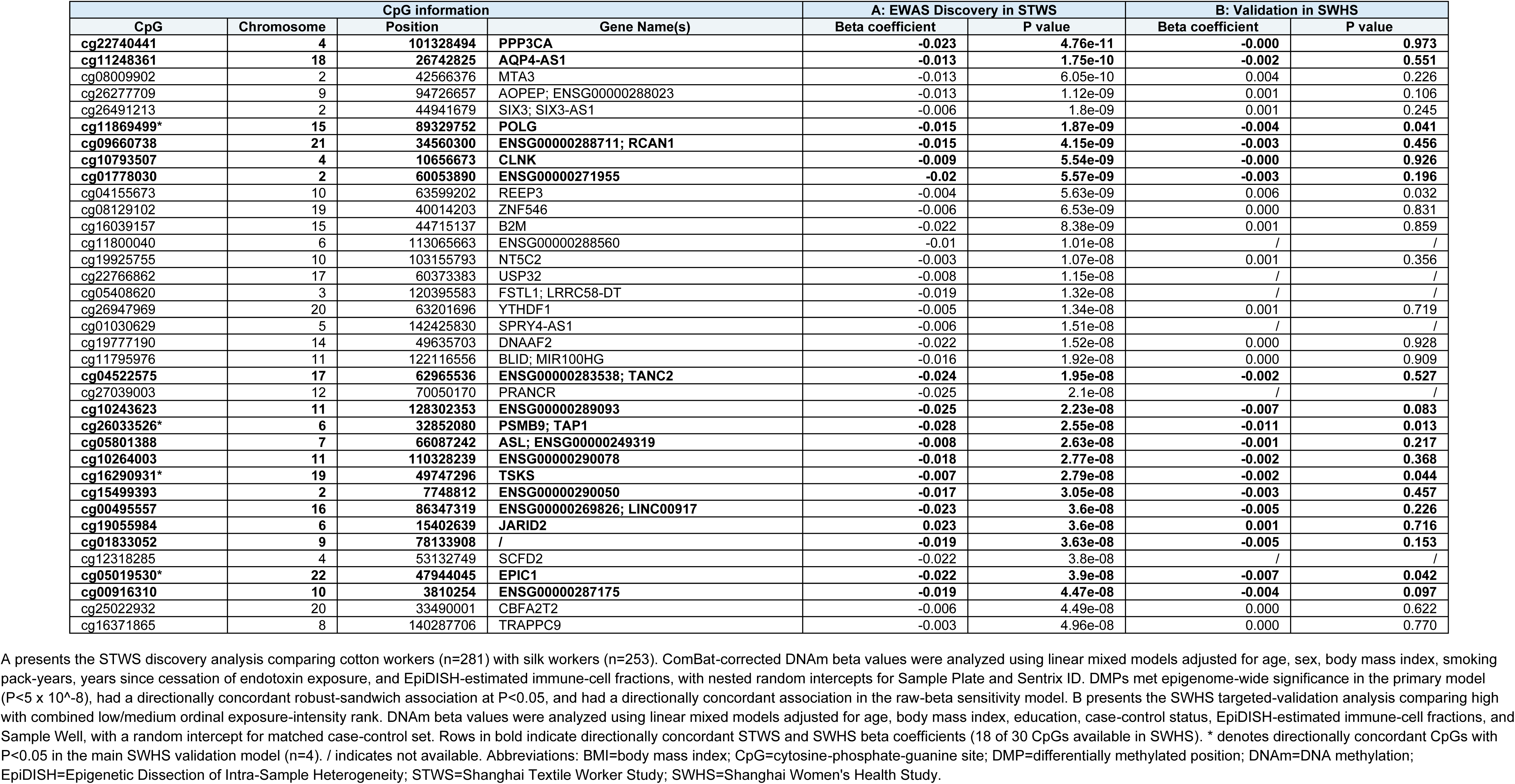
Summary of differentially methylated positions associated with occupational exposure to endotoxin in the Shanghai Textile Worker Study (STWS) and validation in the Shanghai Women’s Health Study (SWHS).

For 15 DMPs, including cg22740441, years since exposure cessation was associated with methylation at P<0.05 in the direction opposite to the endotoxin effect. These results suggest partial reversal of endotoxin-associated hypomethylation after exposure cessation. Supplemental Figure 3 shows adjusted DNAm beta values by cumulative endotoxin exposure and years since exposure cessation.

Compared with silk workers, the low-exposure cotton group showed nominal associations (P<0.05) for 35 of 36 DMPs; the medium- and high-exposure groups showed nominal associations for all 36. Ordered trend tests reached P<0.05 for 34 DMPs. Across all three cotton-exposure groups, the effect direction matched the primary EWAS for each DMP. Much of the exposure-category contrast separated silk workers from workers with any cotton exposure. For 21 DMPs, the low-exposure group had the largest absolute difference from silk workers (Supplemental Table 4).

Cumulative endotoxin was positively associated with all CRP-based IRSs. For each 2.72-fold increase in cumulative endotoxin, associations across the four CRP-based scores ranged from 0.011 SD for IRS_(Ligthart)_ (95% CI, 0.002 to 0.021) to 0.021 SD for IRS_(Hillary-elastic-net)_ (95% CI, 0.011 to 0.031). In contrast, the IL-6-based IRS_(Stevenson)_ was not significantly associated with cumulative endotoxin (Figure 2D). EAA estimates were generally positive, ranging from approximately 1 to 49 additional days per 2.72-fold increase in cumulative endotoxin (Supplemental Figure 4C).

**Figure 2.**
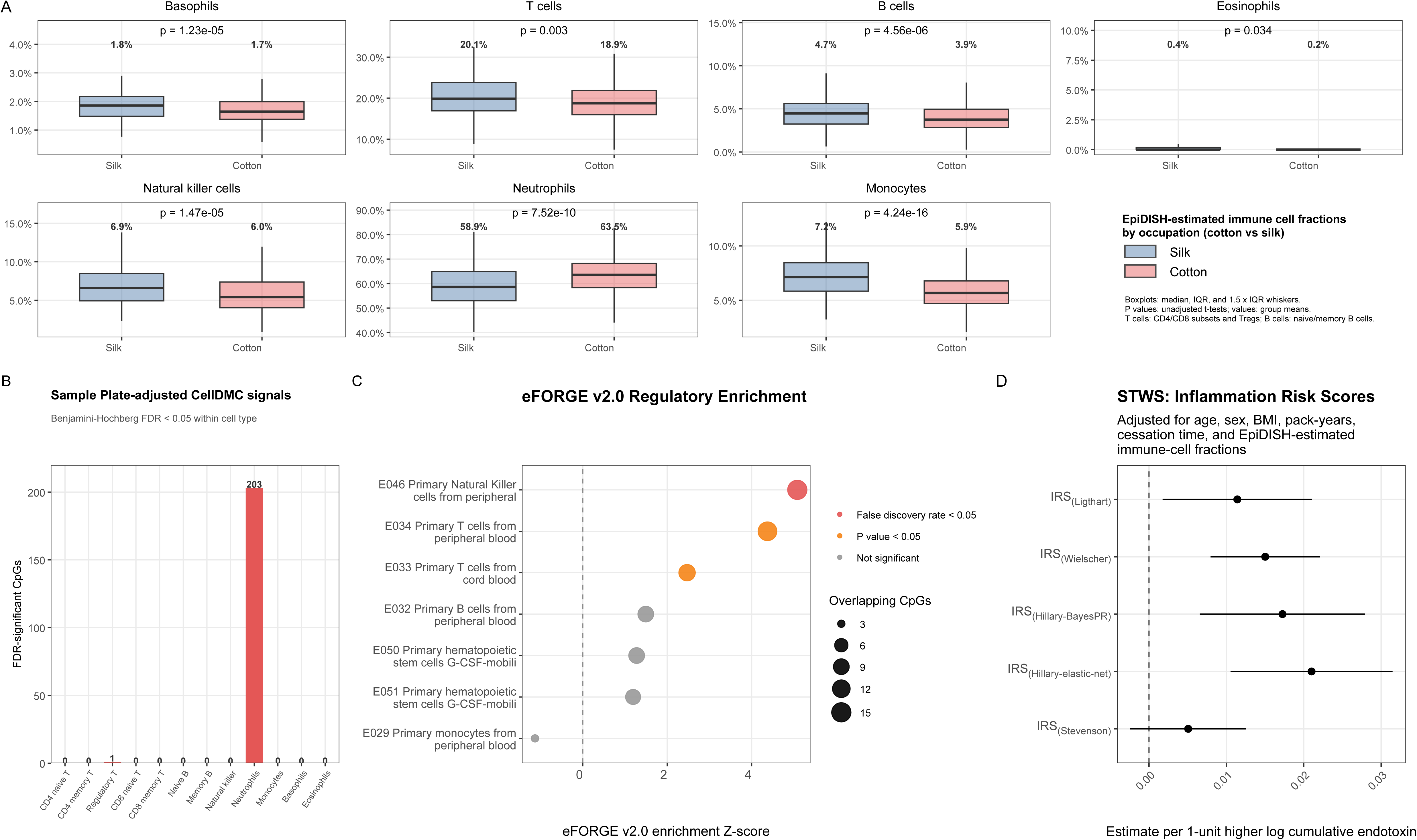
Immune remodeling and inflammation risk in relation to occupational endotoxin exposure. A) Boxplots of EpiDISH-estimated immune cell fractions by occupation (cotton versus silk); P values are from t-tests and numbers above boxes show group means. B) Number of CellDMC signals meeting Benjamini-Hochberg FDR<0.05 within each of 12 immune cell types. CellDMC modeled cotton-versus-silk occupation by EpiDISH-estimated immune-cell fractions and adjusted for age, sex, height, body mass index, smoking pack-years, years since exposure cessation, and Sample Plate. C) eFORGE v2.0 tests whether endotoxin-associated differentially methylated CpGs are enriched in tissue- or cell-type-specific regulatory annotations. D) Associations of cumulative endotoxin with inflammation risk scores in the Shanghai Textile Worker Study from linear models of each standardized score on log cumulative endotoxin, adjusted for age, sex, body mass index, smoking pack-years, years since exposure cessation, and EpiDISH-estimated immune-cell fractions. IRS_(Stevenson)_ is an interleukin-6-based score; the other inflammation risk scores are C-reactive protein-based scores. Abbreviations: BayesPR=Bayesian penalized regression; CellDMC=cell-type-specific differential methylation; CpG=cytosine-phosphate-guanine; EpiDISH=Epigenetic Dissection of Intra-Sample Heterogeneity; eFORGE v2.0=experimentally derived Functional element Overlap analysis of ReGions from Epigenome-Wide Association Studies, version 2.0; FDR=false discovery rate; IRS=inflammation risk score.

#### Targeted Parallel Validation in SWHS

Of the 36 STWS DMPs, 30 were available on the EPIC v1 array used in SWHS. In the primary model comparing high with low or medium exposure-intensity rank, 18 CpGs had concordant effect directions in STWS and SWHS. Four were nominally significant: cg11869499 (*POLG*; P=0.041), cg16290931 (*TSKS*; P=0.044), cg05019530 (*EPIC1*; P=0.042), and cg26033526 (*PSMB9/TAP1*; P=0.013) (Table 2, column B). Supplemental Table 5 reports the sensitivity analyses.

EAA measures showed no clear associations with exposure-intensity rank in SWHS. The CRP-based IRS_(Ligthart)_, IRS_(Wielscher)_, and IRS_(Hillary)_ showed positive associations with high exposure-intensity rank, consistent with the STWS results (Supplemental Figure 4).

### Pathway Enrichment and Immune-Cell Remodeling

#### Pathway enrichment

GSEA identified 152 significant representative gene sets, including 128 GO terms, 10 KEGG pathways, and 14 Reactome pathways (FDR<0.05; Supplemental Figure 5). KEGG and Reactome results centered on immune-system and signal-transduction pathways. The top KEGG pathway was IgSF CAM signaling (hsa04517; 285 genes; FDR=8.20 × 10⁻⁶). The top Reactome pathway was Diseases of signal transduction by growth factor receptors and second messengers (R-HSA-5663202; 450 genes; FDR=3.66 × 10⁻⁷).

Leading-edge analysis identified *ITGB1* and *PIK3CA* as the most recurrent contributors, appearing in 34 and 32 collapsed representative pathways, respectively (Supplemental Table 6). Among genes annotated to endotoxin-associated DMPs, *PPP3CA*, the gene annotated to the most significant DMP (cg22740441), appeared in the most leading-edge pathways. It contributed to six GO biological-process pathways, with the strongest enrichment for positive regulation of cell adhesion (GO:0045785; FDR=0.00248). ORA also highlighted immune and antigen-processing pathways among DMP-mapped genes.

#### Immune-cell remodeling related to endotoxin exposure

EpiDISH, CellDMC, and eFORGE analyses provided complementary evidence of immune-cell remodeling associated with occupational endotoxin exposure. EpiDISH-estimated neutrophil proportions were significantly higher in cotton workers compared with silk workers (63.5% vs. 58.9%; t-test P = 7.52×10^-10^; Figure 2A). Cotton workers also had lower estimated proportions of several lymphoid cell types, including NK cells (5.95% vs. 6.92%; P=1.47 × 10⁻⁵).

CellDMC identified 204 cell-type-specific DNAm signals at FDR<0.05. These signals were observed in neutrophils (n = 203) and regulatory T cells (Treg; n = 1) (Figure 2B). Seven of the 36 DMPs had nominal neutrophil-specific associations at P<0.05.

eFORGE (Figure 2C) found enrichment of the 36 DMPs in blood regulatory regions, including DNase-hypersensitivity sites in primary NK cells from peripheral blood (FDR=0.028, 15 overlapping DMPs). Primary T cells from peripheral blood showed suggestive enrichment (FDR=0.07, 14 DMPs). Collectively, these analyses indicate that cotton workers differed from silk workers in estimated immune-cell composition and cell-type-specific DNAm patterns.

### Targeted respiratory phenotype associations of endotoxin-related DNAm biomarkers

We evaluated 36 endotoxin-associated DMPs and five IRSs in relation to 11 concurrent respiratory phenotypes assessed in 1996 and 40 prospective respiratory phenotypes (Figure 3A and Supplemental Table 7). These targeted associations should be interpreted as exploratory.

**Figure 3.**
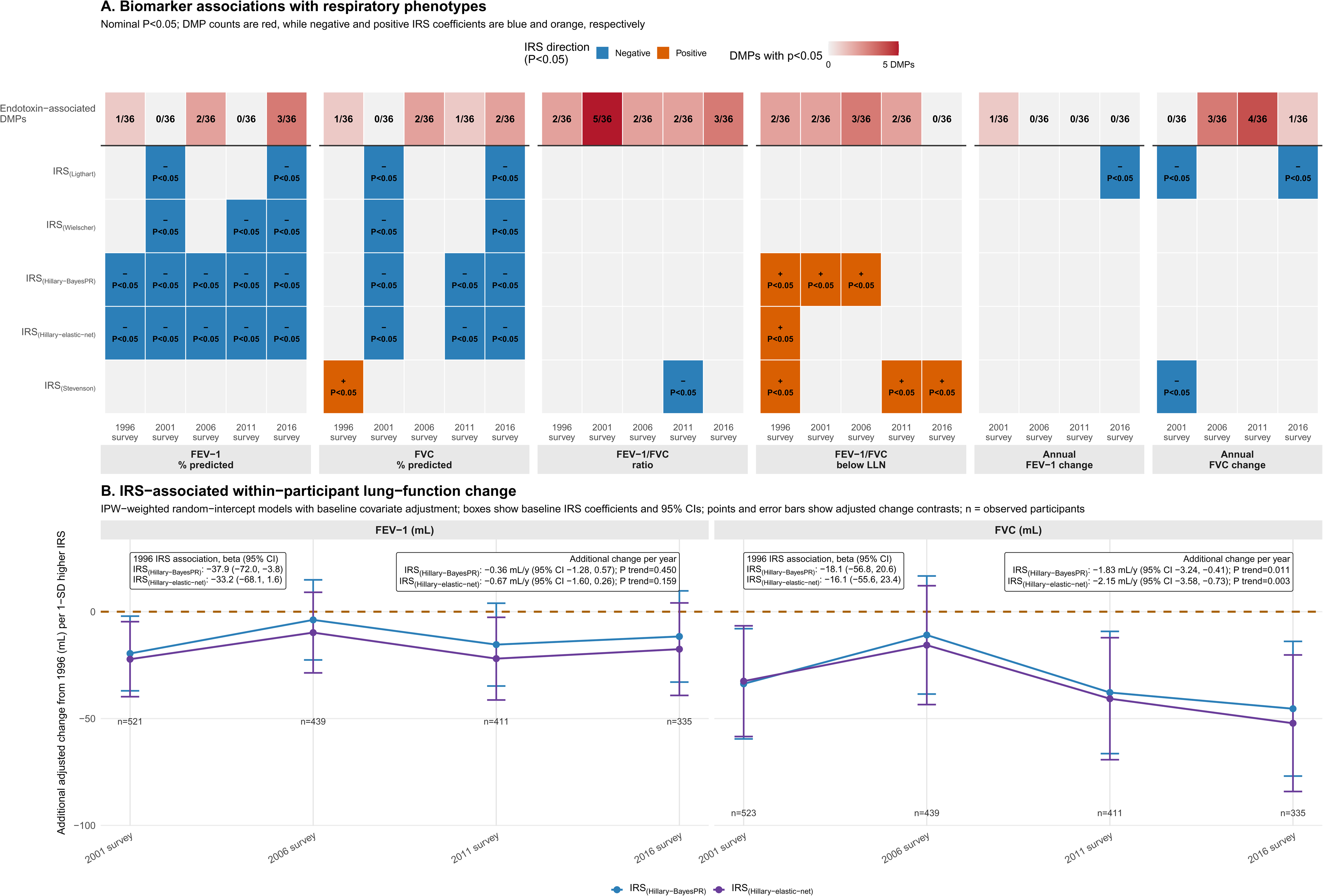
Respiratory phenotype associations with endotoxin-related DNA methylation biomarkers. (A) Heatmap of nominal associations for the 36 endotoxin-associated differentially methylated positions (DMPs) and individual inflammation risk scores (IRSs) with concurrent and prospective respiratory phenotypes; the DMP row gives the number of DMPs with P<0.05. In IRS rows, blue and orange indicate negative and positive model coefficients, respectively; only P<0.05 cells are colored. Colors encode coefficient sign rather than favorable or adverse interpretation, which depends on outcome coding. Separate multivariable linear or logistic regression models adjusted for log cumulative endotoxin, baseline age, sex, height, years since exposure cessation, pack-years, BMI, EpiDISH-estimated immune-cell fractions, and Sample Plate; GLI percent-predicted outcomes omitted age, sex, and height. (B) IPW-weighted random-intercept mixed models, adjusted for the same baseline covariates, estimated the association of 1-SD higher baseline IRS_(Hillary-BayesPR)_ and IRS_(Hillary-elastic-net)_ with FEV-1 and FVC at the 1996 survey and with additional change from 1996 at each subsequent survey; participation weights incorporated survey, prior attendance, baseline IRS, age, sex, pack-years, and cotton work. Boxes give the adjusted 1996 IRS beta coefficients and 95% confidence intervals; error bars are 95% confidence intervals, n denotes observed participants, and labels give the annual IRS-by-time slope, 95% confidence interval, and P trend. Abbreviations: BayesPR, Bayesian penalized regression; DMP, differentially methylated position; FEV-1, forced expiratory volume in 1 second; FVC, forced vital capacity; GLI, Global Lung Function Initiative; IPW, inverse-probability weighting; IRS, inflammation risk score; LLN, lower limit of normal.

Nominal DMP associations occurred most often for airflow and lung-function phenotypes. cg08009902 (*MTA3*) was associated with at least one FEV-1/FVC-based phenotype at every survey after 1996. The top STWS signal, cg22740441 (*PPP3CA*), was associated with percent-predicted FEV-1 in 2016 (3.03% per SD; 95% CI, 0.04 to 6.03). Among the four DMPs nominally validated in SWHS, cg16290931 (*TSKS*) was associated with annual FEV-1 change from 1996 to 2001 (4.46 mL/year per SD; 95% CI, 0.74 to 8.19), and cg05019530 (*EPIC1*) was associated with annual FVC change from 1996 to 2011 (−3.46 mL/year per SD; 95% CI, −6.17 to −0.76).

The CRP-based IRSs showed a more consistent pattern, particularly IRS_(Hillary-BayesPR)_ and IRS_(Hillary-elastic-net)_. Higher scores were associated with lower FEV-1 and FVC at several surveys. The other CRP-based scores had fewer associations. Associations with the IL-6-based IRS_(Stevenson)_ occurred mainly for airflow-obstruction phenotypes in 2011 and 2016.

In the longitudinal analysis, a 1-SD higher 1996 IRS_(Hillary-BayesPR)_ score was associated with 37.9 mL lower FEV-1 in 1996 (95% CI, −72.0 to −3.8). The corresponding estimate for IRS_(Hillary-elastic-net)_ was −33.2 mL (95% CI, −68.1 to 1.6). FEV-1 differences remained negative at later surveys but did not become more negative over follow-up (Ptrend=0.450 and 0.159, respectively). Baseline association between IRS and FVC were insignificant. However, the IRS-associated FVC difference became progressively more negative over time. The additional annual FVC difference was −1.83 mL/year for IRS_(Hillary-BayesPR)_ (95% CI, −3.24 to −0.41; Ptrend=0.011) and −2.15 mL/year for IRS_(Hillary-elastic-net)_ (95% CI, −3.58 to −0.73; Ptrend=0.003). (Figure 3B)

## Discussion

### Overall Summary

In this EWAS of occupational endotoxin exposure, we identified 36 previously unreported endotoxin-associated DNAm sites in STWS, most of which were hypomethylated. Of the 30 signals testable in SWHS, 18 showed directionally concordant associations with occupational exposure-intensity rank, including four that were nominally significant. Analyses of immune-cell composition and cell-type-specific methylation linked cotton work with higher estimated neutrophil proportions and DNAm signals concentrated in neutrophils, regulatory T cells, and lymphoid regulatory regions. Pathway analyses likewise highlighted immune signaling, antigen processing, cell adhesion, and receptor-mediated signaling. Endotoxin was also associated with higher CRP-based DNAm IRSs. In exploratory respiratory analyses, several endotoxin-related DMPs and, more consistently, CRP-based IRSs were associated with lung function and airflow obstruction phenotypes. Higher baseline IRSs were associated with lower FEV-1, while their associations with FVC became progressively more negative over follow-up. Together, these findings suggest that cumulative endotoxin exposure is associated with coordinated DNAm changes reflecting immune remodeling, inflammation, and respiratory impairment.

### Biological relevance of endotoxin-associated DMPs

The top STWS signal, cg22740441 (*PPP3CA*), although not significant in targeted parallel SWHS validation, may still represent a biologically informative endotoxin-associated locus. *PPP3CA* encodes the catalytic subunit of calcineurin, a key component of calcineurin/NFAT signaling in immune cells. Experimental studies of another occupational inhalational exposure, methylene diphenyl diisocyanate, implicate increased *PPP3CA*/calcineurin signaling in inflammatory activation (42), whereas calcineurin signaling is also required for normal fetal lung maturation and surfactant production in mice (43). In STWS, cg22740441 was a potentially reversible DMP, and was associated with current dyspnea and prospective percent predicted FEV-1.

In addition, cg26033526 (*PSMB9/TAP1*), one of the four directionally concordant, nominally significant SWHS loci, has prior associations with HIV infection, inflammatory bowel disease, and eosinophilia (44–46). These observations identify immune-related loci that warrant mechanistic study in the context of occupational endotoxin exposure.

### Endotoxin-related immune remodeling, inflammation risk, and respiratory phenotypes

We observed higher EpiDISH-estimated neutrophil proportions and lower proportions of several lymphoid cell types in cotton workers. CellDMC signals were also concentrated in neutrophils, supporting the cell-composition findings. Elevated blood neutrophil proportions may reflect inflammatory activity linked to airway injury and respiratory impairment (47–49). eFORGE enrichment in NK- and T-cell regulatory regions provided evidence of lymphoid involvement, consistent with studies linking cytotoxic lymphocytes and regulatory T-cell imbalance to lung-function decline (50–52). Together, these findings suggest that long-term endotoxin exposure is associated with coordinated innate inflammatory activation and adaptive immune regulation.

The IRS results provided a complementary measure of inflammation-related DNAm. Cumulative endotoxin was associated with CRP-based DNAm IRSs, whereas the IL-6 score was not significantly associated with endotoxin. This distinction is biologically plausible because IL-6 is an upstream inflammatory cytokine involved in CRP production (53), whereas circulating CRP is a downstream acute-phase protein with distinct production and clearance kinetics (54). In targeted respiratory-phenotype analyses, the CRP-based scores were associated with lower FEV-1 and airflow phenotypes, which is consistent with other population-based cohorts (20).

The convergence of our methylome and prior proteomic findings (9) on immune-related biology supports a role for immune processes in the association between endotoxin exposure and respiratory impairment, with the two omics layers capturing complementary biological time windows. Blood DNAm can provide a relatively stable epigenetic record of past exposure, which is supported by previous EWAS of smoking-associated marks that remained differentially methylated up to decades after cessation and reverted only partially (55). Consistent with this pattern, our endotoxin-associated DMPs likewise tracked years since cessation, suggesting partial reversibility. In contrast, circulating proteins may reflect more proximal inflammatory activity. Within the “meet-in-the-middle” framework (56), analyses integrating exposure, DNAm, proteins, and repeated respiratory measurements could test how these molecular signals relate across time.

### Occupational prevention implications

Despite decades of study, the respiratory literature has not converged on a consensus exposure limit or no-observed-adverse-effect level (NOAEL) for endotoxin. In the Netherlands, an earlier health-based recommendation of 50 EU/m^3^ was later revised by the Dutch Expert Committee on Occupational Safety to 90 EU/m^3^ as an 8-hour time-weighted occupational exposure limit (57). In contrast, experimental cotton-cardroom work suggested no-FEV1-reaction thresholds of approximately 800-1700 EU/m^3^, depending on participant group (58). Previous studies in textile and agricultural workers further suggest that endotoxin effects on respiratory outcomes may be non-linear (6, 59).

The category-specific EWAS results were consistent with a threshold-like pattern at the identified DMPs. Cotton workers in the lowest exposure category (<90 EU/m³) differed from silk workers, while estimates were broadly similar across the low-, medium-, and high-exposure cotton groups. Although they do not establish a biological threshold, these findings reinforce the importance of minimizing endotoxin exposure even at relatively low occupational levels.

### Strengths and Limitations

Our study also had notable strengths. STWS provided detailed historical exposure assessment, decades of follow-up, and repeated respiratory measurements. The EPICv2 array provided broad CpG coverage. Testing STWS signals in an independent cohort reduced reliance on potentially inflated discovery estimates, although differences in population and exposure assessment limited direct replications.

Despite these strengths, several limitations should be considered when interpreting our findings. First, DNAm was measured at a single time point rather than longitudinally. As such, we could not directly track within-person DNAm changes over time. Second, although SWHS provided an independent cohort for targeted parallel validation, endotoxin exposure was assessed using an occupation-derived ordinal exposure-intensity rank. The SWHS rank captured relative occupational exposure potential but not differences in specific tasks, exposure duration, or exposure variability within occupations. Such misclassification may have reduced statistical power and limited direct comparability between the two cohorts. Therefore, the SWHS analysis represents targeted parallel validation using a related exposure proxy rather than an exact replication of the STWS exposure metric. Additionally, SWHS differed from STWS in the distribution of sex, smoking status, and study design. Despite cohort heterogeneity, four loci still showed directionally concordant nominal associations. Finally, healthy-worker survivor effects and differential participation over long follow-up may have influenced observed associations. However, having a healthier study population would likely attenuate findings towards the null and lead to conservative estimation of the true effects.

### Conclusions

Long-term occupational endotoxin exposure was associated with blood DNAm differences, estimated immune-cell remodeling, and enrichment of immune and inflammatory pathways. Endotoxin-related DMPs and CRP-based IRSs also showed exploratory associations with respiratory phenotypes. Independent replication and mechanistic studies are needed to establish the function and clinical relevance of these signals.

## Acknowledgements

We thank all the dedicated staff, past and present, who contributed to the success of our projects. We also extend our appreciation to all the participants of the STWS and SWHS who generously shared their time and made these studies possible.

## Data Availability

STWS data used in this study are available upon reasonable request and subject to approval by the Institutional Review Board of the Harvard T.H. Chan School of Public Health and the Putuo Hospital Institutional Review Board. A data use agreement must be in place prior to data transfer. For inquiries, please contact Dr. David Christiani at.

Access to the SWHS datasets and biospecimens is granted on a case-by-case basis through the SWHS Research Committee via a formalized research application at: https://swhs-smhs.app.vumc.org/forResearchers.php.

Programming code used in the study is available upon request.

## Generative Artificial Intelligence Declaration

Claude Opus 4.8 (Anthropic) and ChatGPT 5.6 (OpenAI) were used for detecting errors and internal inconsistencies, as well as copy editing. The authors take full responsibility for the content of the manuscript.

## Supplemental Tables and Figures Index

- Supplemental Table 1. Shanghai Women’s Health Study occupation-code endotoxin risk dictionary (standalone XLSX; not reproduced in this PDF)
- Supplemental Table 2. Occupational risk of endotoxin in broad occupation groups from the occupation list of Shanghai Women’s Health Study
- Supplemental Table 3. Participant characteristics of the nested case-control cohort of Shanghai Women’s Health Study (SWHS)
- Supplemental Table 4. Annotation and summary of endotoxin-related differentially methylated positions with sensitivity analyses
- Supplemental Table 5. STWS primary and SWHS validation results for 36 differentially methylated positions
- Supplemental Table 6. Gene Set Enrichment Analysis leading-edge genes from collapsed representative pathways
- Supplemental Table 7. Nominally significant biomarker-respiratory outcome associations
- Supplemental Figure 1. Data quality-control workflow
- Supplemental Figure 2. Miami plot
- Supplemental Figure 3. Reversible CpG contour plots
- Supplemental Figure 4. Endotoxin associations with inflammation risk scores and epigenetic accelerated aging
- Supplemental Figure 5. Pathway enrichment

---

**Supplemental Table 2.**
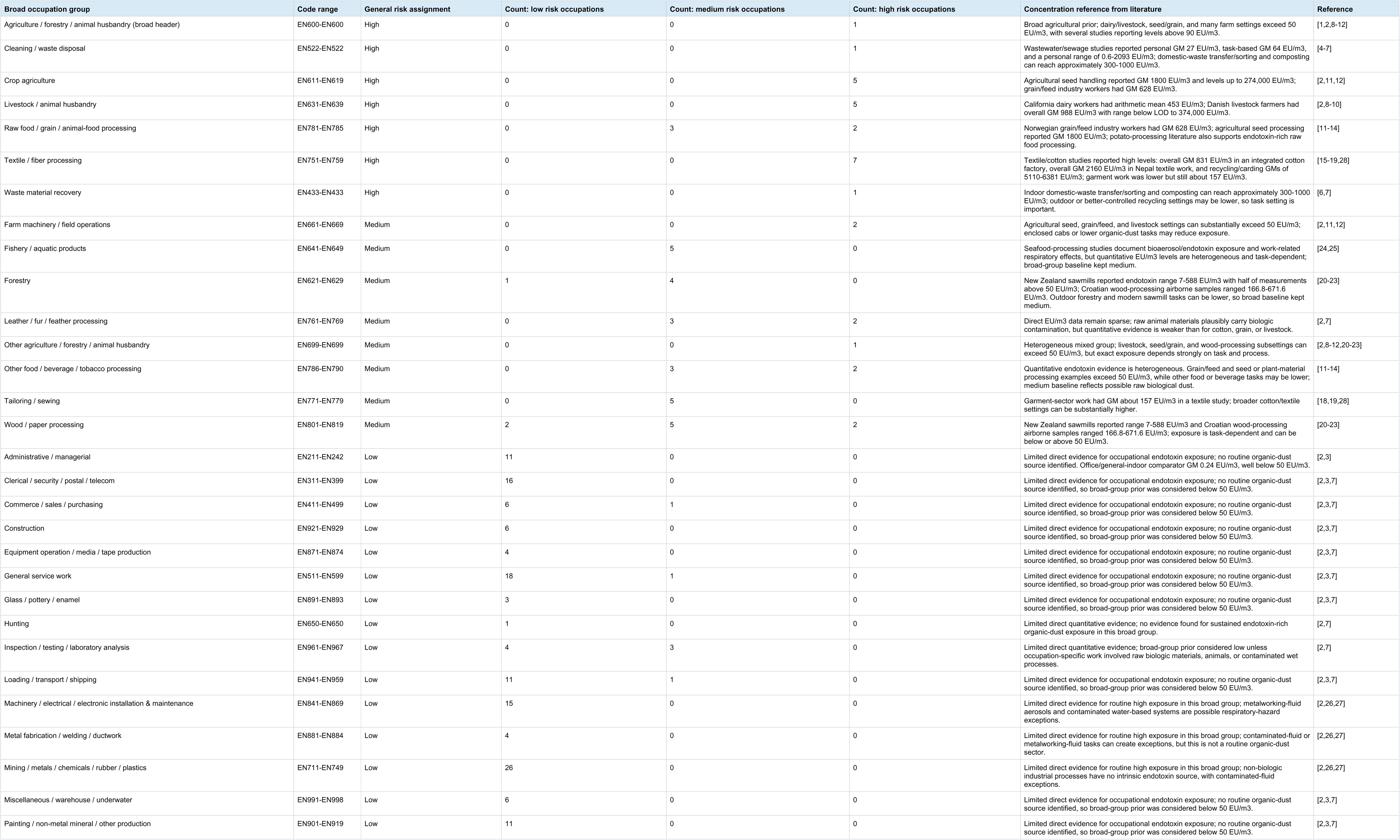
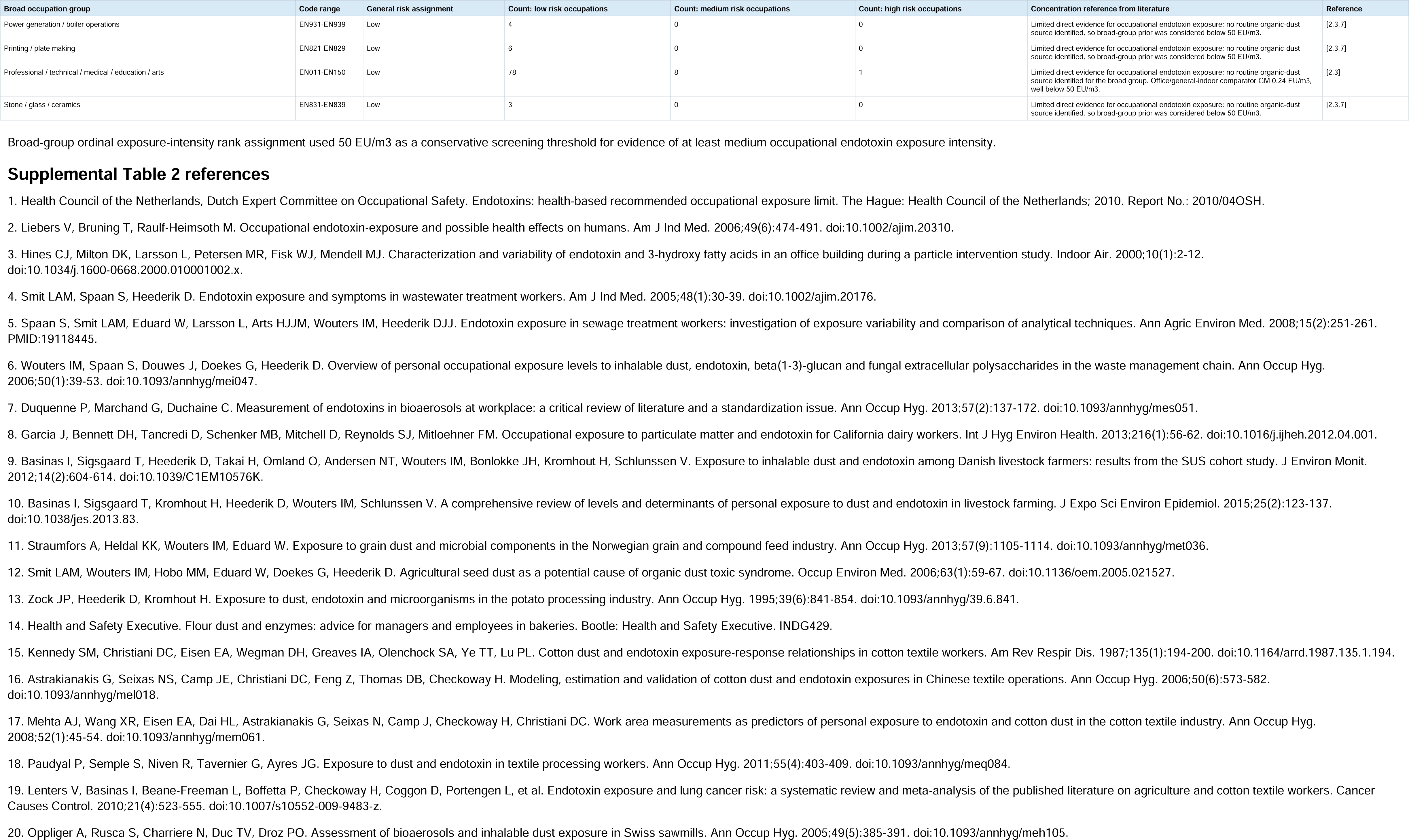
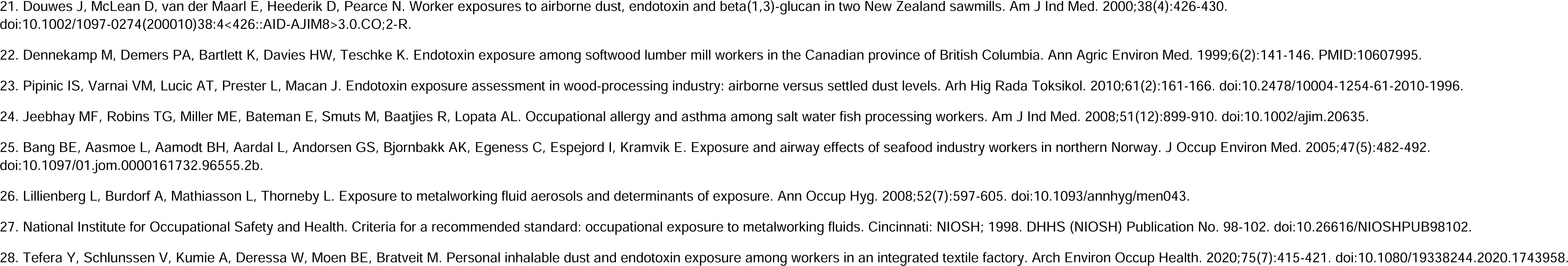
Occupational risk of endotoxin in broad occupation groups from the occupation list of Shanghai Women’s Health Study.

**Supplemental Table 3.**
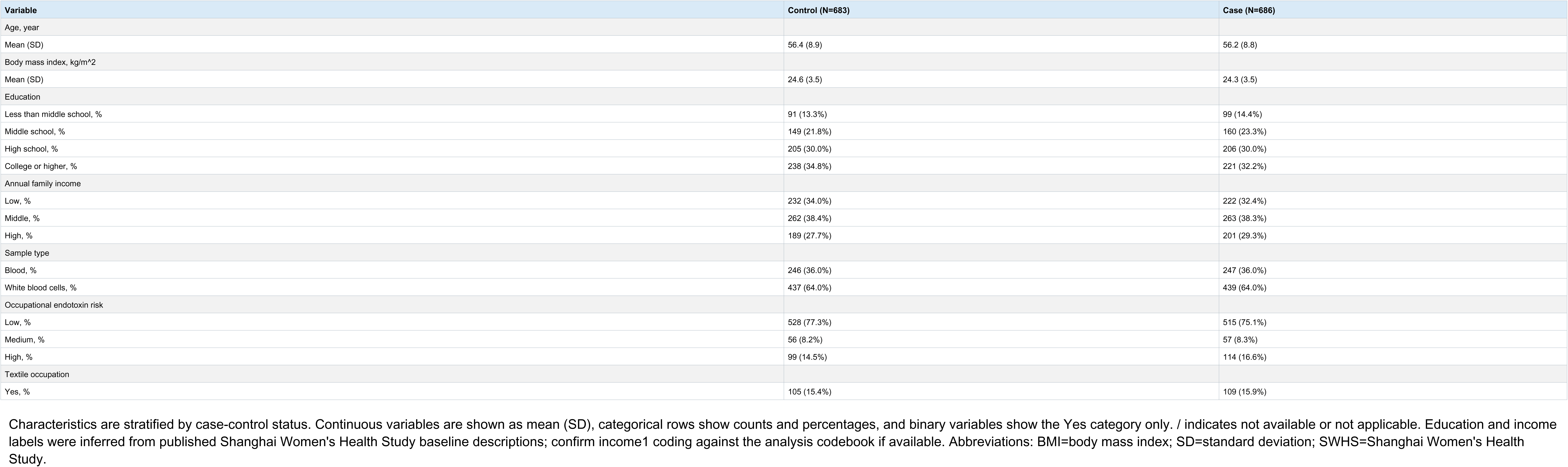
Participant characteristics of the nested case-control cohort of Shanghai Women’s Health Study (SWHS)

**Supplemental Table 4.**
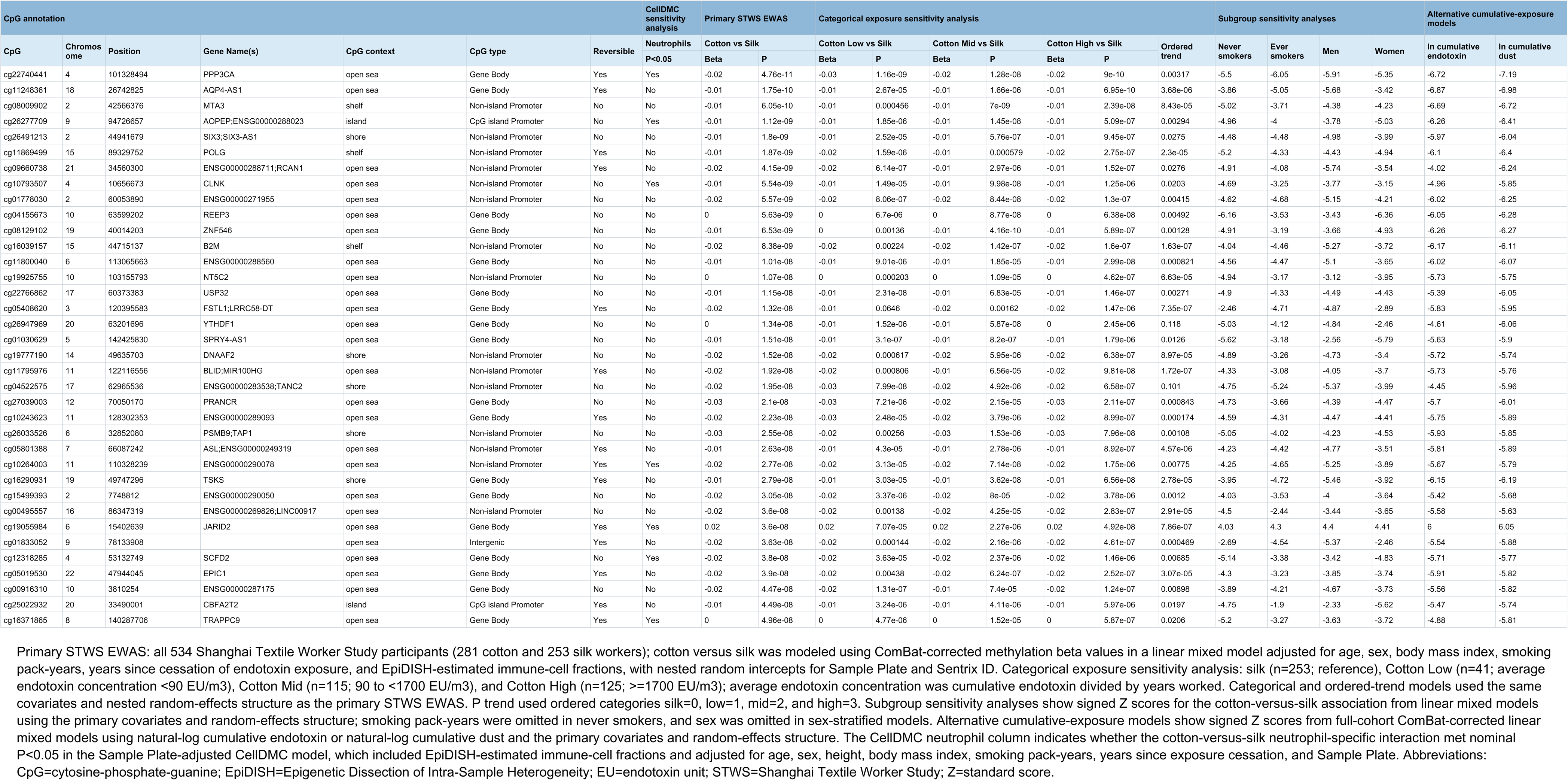
Annotation and summary of endotoxin-related differentially methylated positions with sensitivity analyses.

**Supplemental Table 5.** STWS Primary and SWHS Validation Results for 36 Differentially Methylated Positions.

| CpG | STWS primary |  |  | SWHS primary validation |  |  | SWHS sensitivity validation |  |  |
| --- | --- | --- | --- | --- | --- | --- | --- | --- | --- |
|  | Beta | SE | P value | Beta | SE | P value | Beta | SE | P value |
| cg00495557 | -0.0234 | 0.0042 | 3.60E-08 | -0.0052 | 0.0043 | 0.226 | -0.0063 | 0.0043 | 0.141 |
| cg00916310 | -0.0187 | 0.0032 | 4.47E-08 | -0.0044 | 0.0026 | 0.097 | -0.0020 | 0.0027 | 0.441 |
| cg01030629 | -0.0058 | 0.0010 | 1.51E-08 | / | / | / | / | / | / |
| cg01778030 | -0.0201 | 0.0032 | 5.57E-09 | -0.0031 | 0.0024 | 0.196 | -0.0023 | 0.0024 | 0.351 |
| cg01833052 | -0.0191 | 0.0033 | 3.63E-08 | -0.0046 | 0.0032 | 0.153 | -0.0024 | 0.0034 | 0.479 |
| cg04155673 | -0.0040 | 0.0006 | 5.63E-09 | 0.0060 | 0.0028 | 0.032 | 0.0065 | 0.0029 | 0.023 |
| cg04522575 | -0.0239 | 0.0040 | 1.95E-08 | -0.0022 | 0.0035 | 0.527 | 0.0028 | 0.0035 | 0.415 |
| cg05019530 | -0.0223 | 0.0038 | 3.90E-08 | -0.0074 | 0.0036 | 0.042 | -0.0053 | 0.0037 | 0.148 |
| cg05408620 | -0.0194 | 0.0034 | 1.32E-08 | / | / | / | / | / | / |
| cg05801388 | -0.0078 | 0.0013 | 2.63E-08 | -0.0013 | 0.0010 | 0.217 | -0.0004 | 0.0011 | 0.726 |
| cg08009902 | -0.0130 | 0.0020 | 6.05E-10 | 0.0038 | 0.0031 | 0.226 | 0.0059 | 0.0032 | 0.065 |
| cg08129102 | -0.0059 | 0.0010 | 6.53E-09 | 0.0005 | 0.0023 | 0.831 | -0.0001 | 0.0024 | 0.978 |
| cg09660738 | -0.0152 | 0.0024 | 4.15E-09 | -0.0030 | 0.0041 | 0.456 | -0.0037 | 0.0041 | 0.374 |
| cg10243623 | -0.0249 | 0.0042 | 2.23E-08 | -0.0066 | 0.0038 | 0.084 | -0.0033 | 0.0038 | 0.382 |
| cg10264003 | -0.0184 | 0.0031 | 2.77E-08 | -0.0022 | 0.0024 | 0.368 | -0.0014 | 0.0025 | 0.564 |
| cg10793507 | -0.0095 | 0.0015 | 5.54E-09 | -0.0004 | 0.0042 | 0.926 | 0.0019 | 0.0044 | 0.654 |
| cg11248361 | -0.0131 | 0.0019 | 1.75E-10 | -0.0018 | 0.0029 | 0.551 | -0.0022 | 0.0030 | 0.456 |
| cg11795976 | -0.0160 | 0.0028 | 1.92E-08 | 0.0005 | 0.0040 | 0.909 | 0.0018 | 0.0042 | 0.662 |
| cg11800040 | -0.0101 | 0.0017 | 1.01E-08 | / | / | / | / | / | / |
| cg11869499 | -0.0146 | 0.0023 | 1.87E-09 | -0.0044 | 0.0022 | 0.041 | -0.0002 | 0.0022 | 0.929 |
| cg12318285 | -0.0220 | 0.0038 | 3.80E-08 | / | / | / | / | / | / |
| cg15499393 | -0.0172 | 0.0031 | 3.05E-08 | -0.0028 | 0.0038 | 0.457 | 0.0006 | 0.0039 | 0.878 |
| cg16039157 | -0.0215 | 0.0035 | 8.38E-09 | 0.0006 | 0.0037 | 0.859 | 0.0048 | 0.0036 | 0.184 |
| cg16290931 | -0.0074 | 0.0012 | 2.79E-08 | -0.0016 | 0.0008 | 0.044 | -0.0015 | 0.0008 | 0.062 |
| cg16371865 | -0.0033 | 0.0006 | 4.96E-08 | 0.0005 | 0.0016 | 0.770 | 0.0006 | 0.0016 | 0.696 |
| cg19055984 | 0.0227 | 0.0038 | 3.60E-08 | 0.0012 | 0.0034 | 0.716 | 0.0013 | 0.0034 | 0.707 |
| cg19777190 | -0.0219 | 0.0038 | 1.52E-08 | 0.0003 | 0.0032 | 0.928 | -0.0008 | 0.0032 | 0.810 |
| cg19925755 | -0.0033 | 0.0006 | 1.07E-08 | 0.0011 | 0.0012 | 0.356 | 0.0004 | 0.0012 | 0.741 |
| cg22740441 | -0.0225 | 0.0031 | 4.76E-11 | -0.0001 | 0.0033 | 0.973 | 0.0058 | 0.0034 | 0.090 |
| cg22766862 | -0.0077 | 0.0013 | 1.15E-08 | / | / | / | / | / | / |
| cg25022932 | -0.0059 | 0.0010 | 4.49E-08 | 0.0004 | 0.0009 | 0.622 | 0.0009 | 0.0009 | 0.310 |
| cg26033526 | -0.0277 | 0.0047 | 2.55E-08 | -0.0110 | 0.0044 | 0.013 | -0.0049 | 0.0045 | 0.273 |
| cg26277709 | -0.0129 | 0.0020 | 1.12E-09 | 0.0007 | 0.0004 | 0.106 | 0.0005 | 0.0005 | 0.276 |
| cg26491213 | -0.0064 | 0.0010 | 1.80E-09 | 0.0007 | 0.0006 | 0.245 | 0.0007 | 0.0006 | 0.213 |
| cg26947969 | -0.0049 | 0.0008 | 1.34E-08 | 0.0010 | 0.0027 | 0.719 | 0.0019 | 0.0027 | 0.483 |
| cg27039003 | -0.0251 | 0.0042 | 2.10E-08 | / | / | / | / | / | / |

**STWS primary:** ComBat-corrected DNAm beta values were compared between cotton and silk workers using linear mixed regression adjusted for age, sex, body mass index, smoking pack-years, years since cessation of endotoxin exposure, and EpiDISH-estimated immune-cell fractions, with nested random intercepts for Sample Plate and Sentrix ID.
**SWHS primary validation:** women with high ordinal exposure-intensity rank were compared with women in the combined low/medium group using linear mixed regression adjusted for age, body mass index, education, case-control status, EpiDISH-estimated immune-cell fractions, and Sample Well, with a random intercept for matched case-control set.
**SWHS sensitivity validation:** the same high versus combined low/medium comparison used the same covariates and random intercept and was weighted using inverse-probability weights for case-control status.
**Abbreviations:** BMI=body mass index; CpG=cytosine-phosphate-guanine; DMP=differentially methylated position; EpiDISH=Epigenetic Dissection of Intra-Sample Heterogeneity; IPW=inverse-probability weighting; SE=standard error; STWS=Shanghai Textile Worker Study; SWHS=Shanghai Women’s Health Study. / indicates a CpG unavailable in SWHS.

**Supplemental Table 6.**
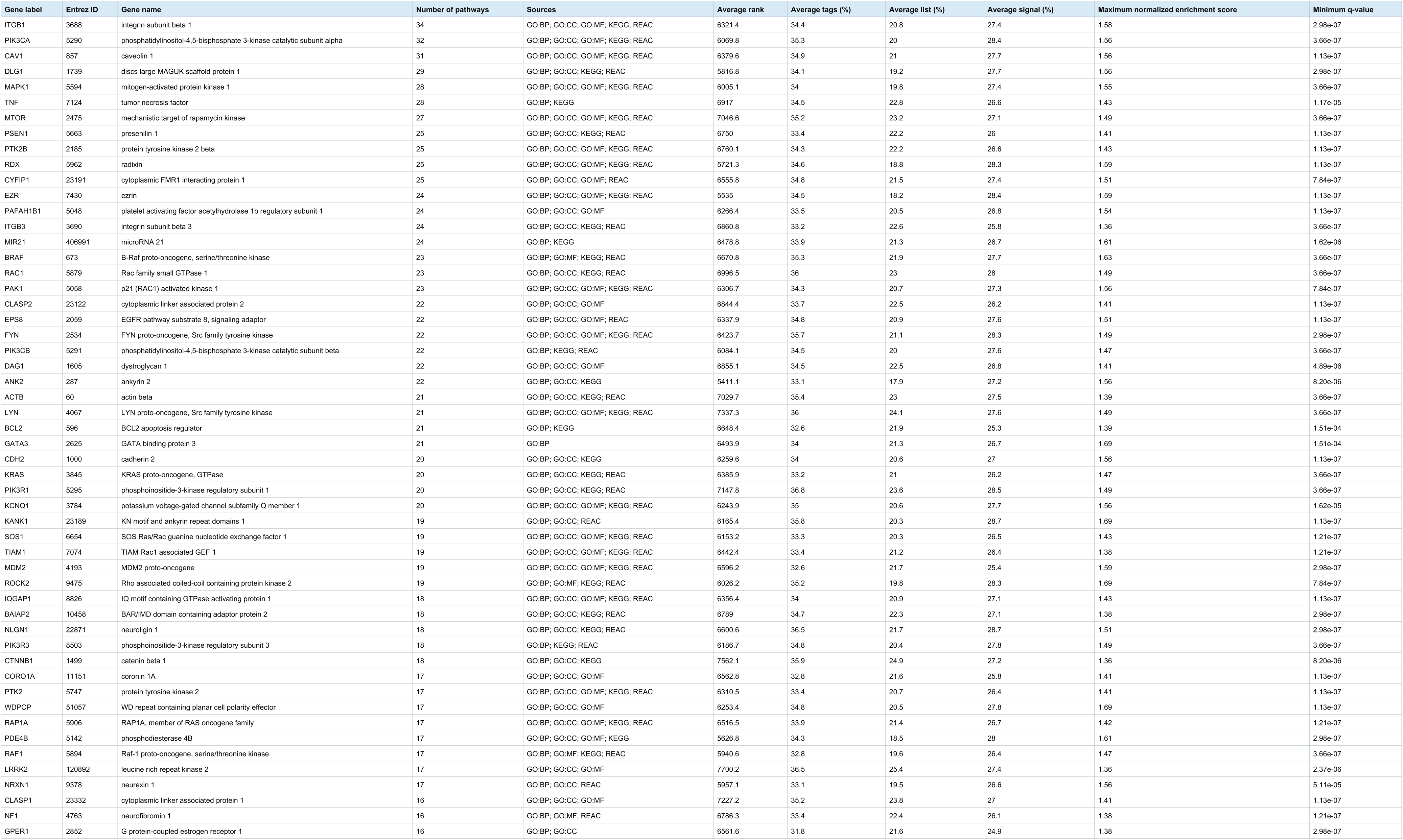
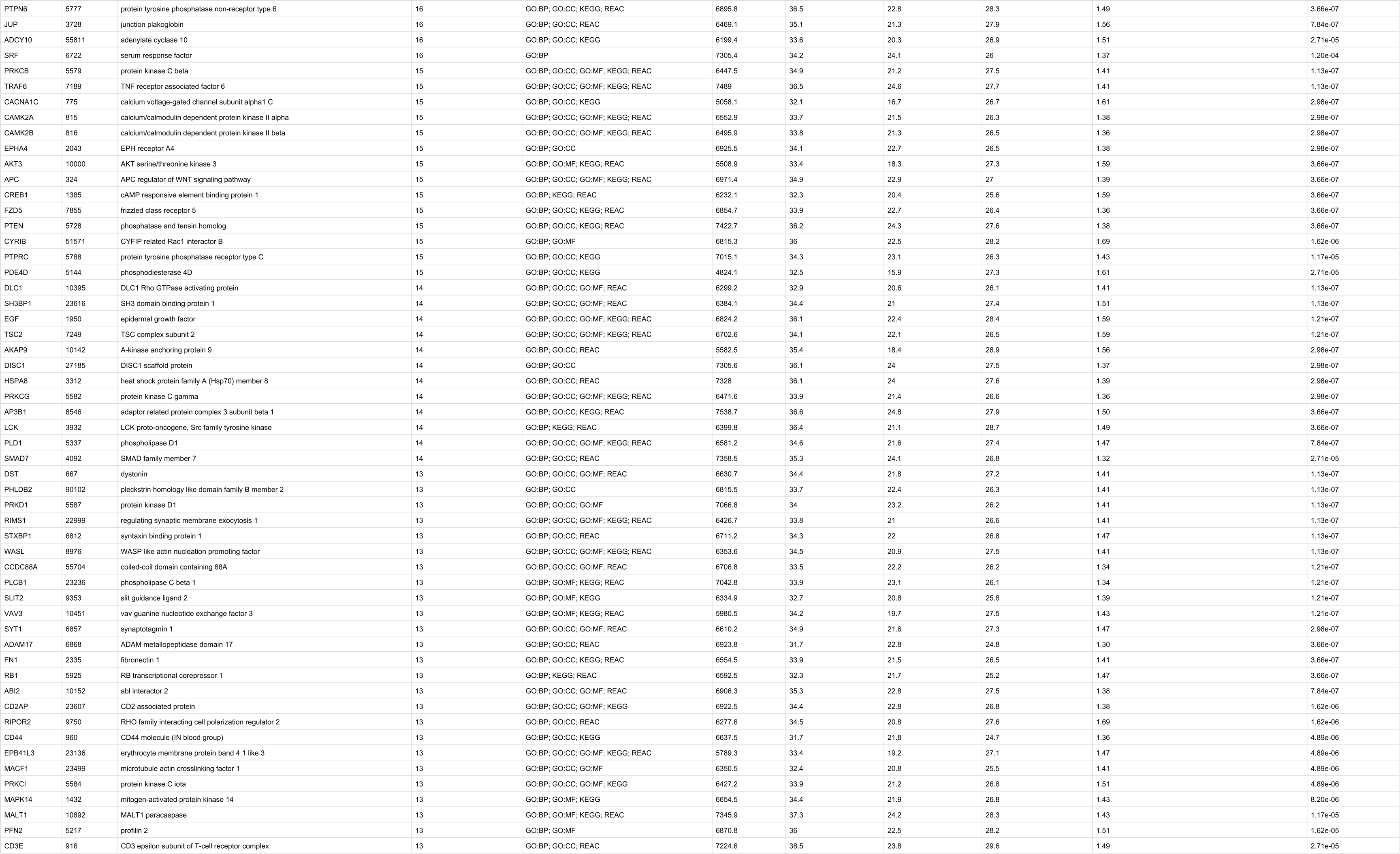
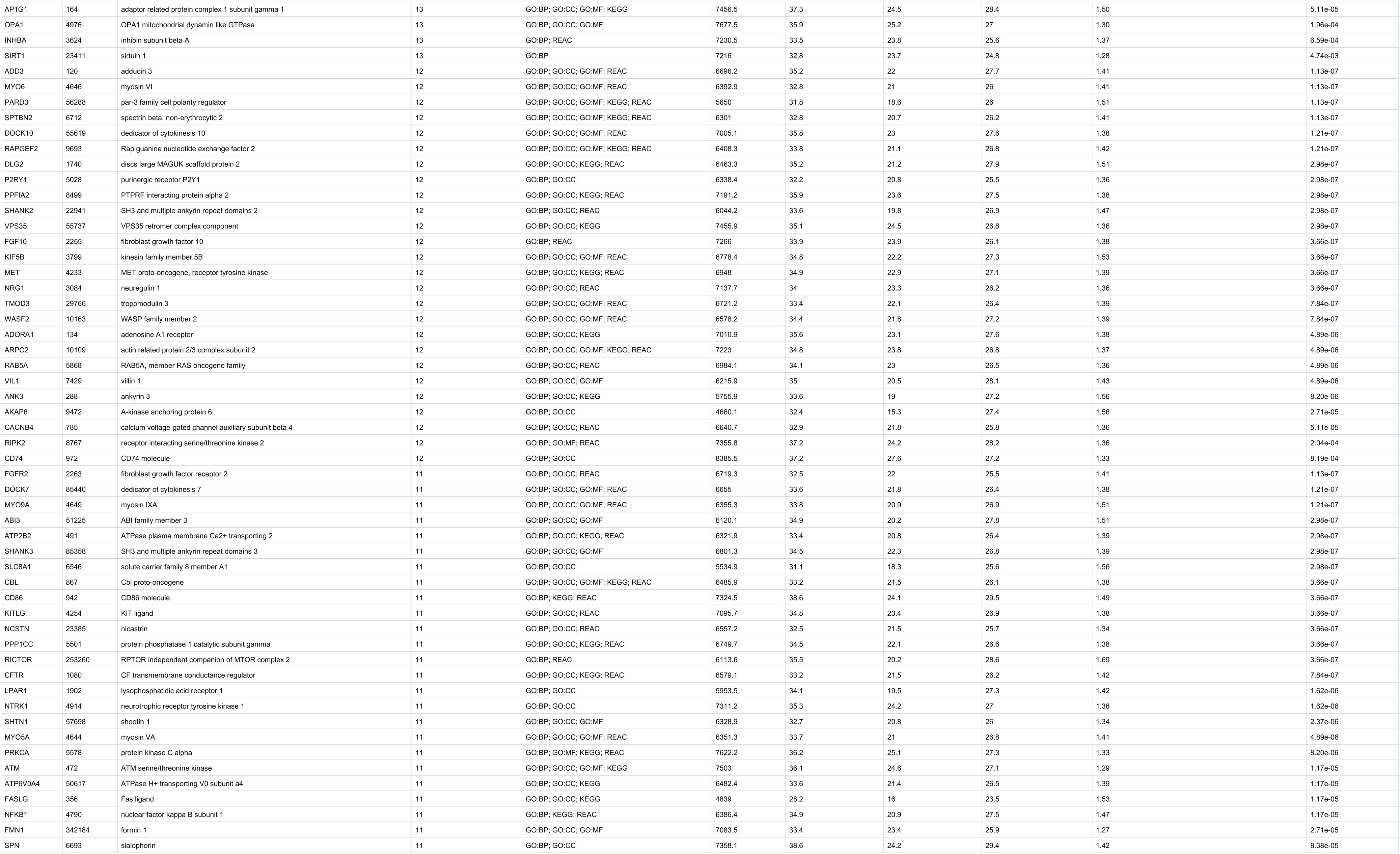
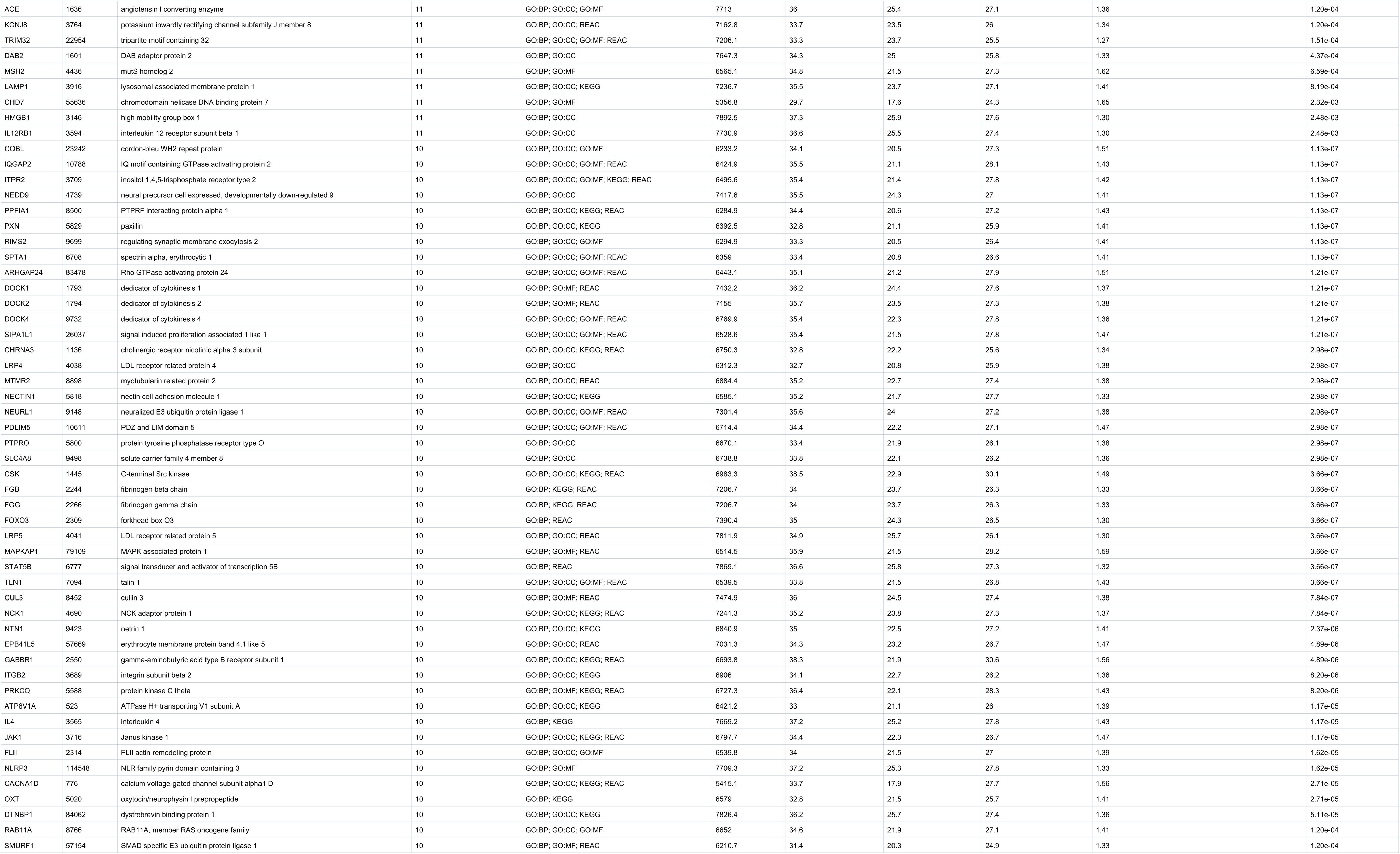
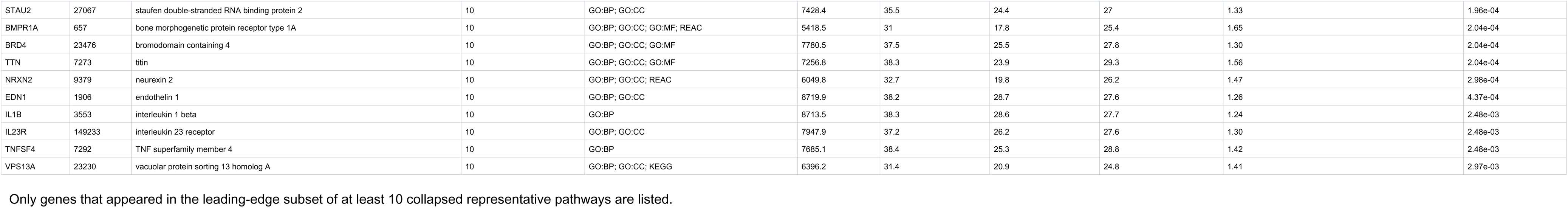
Gene Set Enrichment Analysis leading-edge genes from collapsed representative pathways.

**Supplemental Table 7.** Nominally Significant Biomarker-Respiratory Outcome Associations.

| Analysis Timing | Biomarker | Biomarker Type | Outcome | Effect scale | Effect | Beta / OR | SE (Beta only) | P | 95% CI |
| --- | --- | --- | --- | --- | --- | --- | --- | --- | --- |
| Concurrent (1996 survey) | cg00916310 | DMPs | Dyspnea | 1-SD higher methylation beta | OR | 1.771 |  | 1.3500E-02 | [1.125, 2.788] |
| Concurrent (1996 survey) | cg01778030 | DMPs | Dyspnea | 1-SD higher methylation beta | OR | 1.882 |  | 1.8253E-02 | [1.113, 3.182] |
| Concurrent (1996 survey) | cg01778030 | DMPs | FVC % predicted | 1-SD higher methylation beta | Beta | 2.006 | 0.877 | 2.2590E-02 | [0.287, 3.725] |
| Concurrent (1996 survey) | cg01778030 | DMPs | FVC (mL) | 1-SD higher methylation beta | Beta | 63.956 | 32.372 | 4.8768E-02 | [0.507, 127.404] |
| Concurrent (1996 survey) | cg01833052 | DMPs | Dyspnea | 1-SD higher methylation beta | OR | 1.744 |  | 4.0467E-02 | [1.024, 2.969] |
| Concurrent (1996 survey) | cg08009902 | DMPs | FEV-1/FVC <0.70 | 1-SD higher methylation beta | OR | 0.728 |  | 4.2695E-02 | [0.536, 0.990] |
| Concurrent (1996 survey) | cg08129102 | DMPs | FEV-1/FVC ratio | 1-SD higher methylation beta | Beta | -0.008 | 0.004 | 3.6477E-02 | [-0.015, -0.001] |
| Concurrent (1996 survey) | cg11248361 | DMPs | FEV-1/FVC ratio | 1-SD higher methylation beta | Beta | 0.009 | 0.004 | 2.7731E-02 | [0.001, 0.017] |
| Concurrent (1996 survey) | cg11795976 | DMPs | Dyspnea | 1-SD higher methylation beta | OR | 1.803 |  | 3.0975E-02 | [1.055, 3.080] |
| Concurrent (1996 survey) | cg16290931 | DMPs | Dyspnea | 1-SD higher methylation beta | OR | 1.559 |  | 4.2444E-02 | [1.015, 2.395] |
| Concurrent (1996 survey) | cg16371865 | DMPs | Chronic cough | 1-SD higher methylation beta | OR | 2.151 |  | 3.8329E-02 | [1.042, 4.442] |
| Concurrent (1996 survey) | cg19055984 | DMPs | FEV-1/FVC < GLI LLN | 1-SD higher methylation beta | OR | 1.457 |  | 2.3074E-02 | [1.053, 2.015] |
| Concurrent (1996 survey) | cg19777190 | DMPs | Chronic bronchitis | 1-SD higher methylation beta | OR | 0.414 |  | 1.4248E-02 | [0.204, 0.838] |
| Concurrent (1996 survey) | cg19777190 | DMPs | Chronic cough | 1-SD higher methylation beta | OR | 0.454 |  | 4.3479E-02 | [0.211, 0.977] |
| Concurrent (1996 survey) | cg22740441 | DMPs | Dyspnea | 1-SD higher methylation beta | OR | 1.724 |  | 4.2204E-02 | [1.019, 2.915] |
| Concurrent (1996 survey) | cg26033526 | DMPs | Dyspnea | 1-SD higher methylation beta | OR | 1.984 |  | 7.4708E-03 | [1.201, 3.277] |
| Concurrent (1996 survey) | cg26491213 | DMPs | FEV-1/FVC <0.70 | 1-SD higher methylation beta | OR | 0.660 |  | 5.3918E-03 | [0.493, 0.884] |
| Concurrent (1996 survey) | cg26491213 | DMPs | FEV-1/FVC < GLI LLN | 1-SD higher methylation beta | OR | 0.690 |  | 1.9398E-02 | [0.506, 0.942] |
| Concurrent (1996 survey) | cg26491213 | DMPs | FEV-1 % predicted | 1-SD higher methylation beta | Beta | 1.426 | 0.638 | 2.5869E-02 | [0.176, 2.677] |
| Concurrent (1996 survey) | cg26947969 | DMPs | Dyspnea | 1-SD higher methylation beta | OR | 1.449 |  | 4.3993E-02 | [1.010, 2.078] |
| Concurrent (1996 survey) | IRS<sub>(Hillary-BayesPR)</sub> | IRS | FEV-1/FVC <0.70 | 1-SD higher score | OR | 1.581 |  | 1.6193E-03 | [1.189, 2.103] |
| Concurrent (1996 survey) | IRS<sub>(Hillary-BayesPR)</sub> | IRS | FEV-1/FVC < GLI LLN | 1-SD higher score | OR | 1.573 |  | 3.1269E-03 | [1.165, 2.125] |
| Concurrent (1996 survey) | IRS<sub>(Hillary-BayesPR)</sub> | IRS | FEV-1 (mL) | 1-SD higher score | Beta | -42.722 | 18.167 | 1.9099E-02 | [-78.329, -7.115] |
| Concurrent (1996 survey) | IRS<sub>(Hillary-BayesPR)</sub> | IRS | FEV-1 % predicted | 1-SD higher score | Beta | -1.410 | 0.623 | 2.4116E-02 | [-2.631, -0.188] |
| Concurrent (1996 survey) | IRS<sub>(Hillary-elastic-net)</sub> | IRS | FEV-1/FVC < GLI LLN | 1-SD higher score | OR | 1.522 |  | 7.5943E-03 | [1.118, 2.072] |
| Concurrent (1996 survey) | IRS<sub>(Hillary-elastic-net)</sub> | IRS | FEV-1/FVC <0.70 | 1-SD higher score | OR | 1.466 |  | 9.4640E-03 | [1.098, 1.956] |
| Concurrent (1996 survey) | IRS<sub>(Hillary-elastic-net)</sub> | IRS | FEV-1 (mL) | 1-SD higher score | Beta | -40.087 | 18.647 | 3.2077E-02 | [-76.634, -3.539] |
| Concurrent (1996 survey) | IRS<sub>(Hillary-elastic-net)</sub> | IRS | FEV-1 % predicted | 1-SD higher score | Beta | -1.295 | 0.639 | 4.3369E-02 | [-2.548, -0.042] |
| Concurrent (1996 survey) | IRS<sub>(Stevenson)</sub> | IRS | FVC % predicted | 1-SD higher score | Beta | 2.048 | 0.770 | 8.0628E-03 | [0.539, 3.557] |
| Concurrent (1996 survey) | IRS<sub>(Stevenson)</sub> | IRS | FEV-1/FVC < GLI LLN | 1-SD higher score | OR | 1.614 |  | 3.4772E-02 | [1.035, 2.517] |
| Prospective | cg01778030 | DMPs | Chronic cough 2001 | 1-SD higher methylation beta | OR | 1.832 |  | 4.5705E-02 | [1.012, 3.319] |
| Prospective | cg04522575 | DMPs | Annual FVC change 1996–2006 | 1-SD higher methylation beta | Beta | -5.137 | 1.926 | 7.9654E-03 | [-8.913, -1.362] |
| Prospective | cg04522575 | DMPs | Annual FVC change 1996–2011 | 1-SD higher methylation beta | Beta | -3.225 | 1.277 | 1.1962E-02 | [-5.728, -0.723] |
| Prospective | cg05019530 | DMPs | Annual FVC change 1996–2011 | 1-SD higher methylation beta | Beta | -3.461 | 1.380 | 1.2541E-02 | [-6.165, -0.757] |
| Prospective | cg05408620 | DMPs | Chronic bronchitis 2001 | 1-SD higher methylation beta | OR | 1.978 |  | 2.9454E-02 | [1.070, 3.657] |
| Prospective | cg05801388 | DMPs | Annual FVC change 1996–2016 | 1-SD higher methylation beta | Beta | 1.841 | 0.896 | 4.0818E-02 | [0.084, 3.597] |
| Prospective | cg05801388 | DMPs | FEV-1/FVC ratio 2001 | 1-SD higher methylation beta | Beta | 0.007 | 0.003 | 4.5907E-02 | [0.000, 0.013] |
| Prospective | cg08009902 | DMPs | FEV-1/FVC ratio 2016 | 1-SD higher methylation beta | Beta | 0.010 | 0.004 | 8.3516E-03 | [0.003, 0.018] |
| Prospective | cg08009902 | DMPs | FEV-1/FVC < GLI LLN 2001 | 1-SD higher methylation beta | OR | 0.535 |  | 9.0123E-03 | [0.335, 0.856] |
| Prospective | cg08009902 | DMPs | FEV-1/FVC < GLI LLN 2006 | 1-SD higher methylation beta | OR | 0.584 |  | 1.8344E-02 | [0.374, 0.913] |
| Prospective | cg08009902 | DMPs | FEV-1/FVC < GLI LLN 2011 | 1-SD higher methylation beta | OR | 0.530 |  | 2.7134E-02 | [0.302, 0.931] |
| Prospective | cg08009902 | DMPs | FEV-1/FVC <0.70 2011 | 1-SD higher methylation beta | OR | 0.658 |  | 3.5551E-02 | [0.445, 0.972] |
| Prospective | cg08009902 | DMPs | FEV-1/FVC ratio 2006 | 1-SD higher methylation beta | Beta | 0.008 | 0.004 | 3.7471E-02 | [0.001, 0.016] |
| Prospective | cg08009902 | DMPs | FEV-1/FVC ratio 2011 | 1-SD higher methylation beta | Beta | 0.008 | 0.004 | 4.3771E-02 | [0.000, 0.015] |
| Prospective | cg08009902 | DMPs | FEV-1/FVC <0.70 2001 | 1-SD higher methylation beta | OR | 0.686 |  | 4.9336E-02 | [0.471, 0.999] |
| Prospective | cg08129102 | DMPs | FEV-1/FVC ratio 2001 | 1-SD higher methylation beta | Beta | -0.007 | 0.003 | 3.1917E-02 | [-0.014, -0.001] |
| Prospective | cg10264003 | DMPs | Annual FVC change 1996–2006 | 1-SD higher methylation beta | Beta | -5.494 | 2.352 | 1.9970E-02 | [-10.103, -0.885] |
| Prospective | cg10793507 | DMPs | FVC (mL) 2011 | 1-SD higher methylation beta | Beta | -88.826 | 26.702 | 9.6192E-04 | [-141.161, -36.490] |
| Prospective | cg10793507 | DMPs | FVC % predicted 2011 | 1-SD higher methylation beta | Beta | -2.623 | 0.799 | 1.1222E-03 | [-4.190, -1.057] |
| Prospective | cg10793507 | DMPs | Annual FVC change 1996–2011 | 1-SD higher methylation beta | Beta | -3.531 | 1.135 | 2.0130E-03 | [-5.757, -1.306] |
| Prospective | cg10793507 | DMPs | FEV-1/FVC <0.70 2011 | 1-SD higher methylation beta | OR | 0.587 |  | 5.9906E-03 | [0.402, 0.858] |
| Prospective | cg10793507 | DMPs | FVC % predicted 2006 | 1-SD higher methylation beta | Beta | -1.900 | 0.702 | 7.0604E-03 | [-3.276, -0.524] |
| Prospective | cg10793507 | DMPs | FVC (mL) 2006 | 1-SD higher methylation beta | Beta | -63.520 | 24.499 | 9.8539E-03 | [-111.538, -15.501] |
| Prospective | cg10793507 | DMPs | FVC (mL) 2016 | 1-SD higher methylation beta | Beta | -67.709 | 30.346 | 2.6365E-02 | [-127.188, -8.231] |
| Prospective | cg10793507 | DMPs | FEV-1/FVC < GLI LLN 2011 | 1-SD higher methylation beta | OR | 0.559 |  | 2.9161E-02 | [0.332, 0.943] |
| Prospective | cg10793507 | DMPs | FVC % predicted 2016 | 1-SD higher methylation beta | Beta | -2.081 | 1.001 | 3.8446E-02 | [-4.044, -0.119] |
| Prospective | cg10793507 | DMPs | FEV-1/FVC ratio 2011 | 1-SD higher methylation beta | Beta | 0.008 | 0.004 | 3.8683E-02 | [0.000, 0.016] |
| Prospective | cg11248361 | DMPs | FEV-1/FVC <0.70 2011 | 1-SD higher methylation beta | OR | 0.619 |  | 2.4163E-02 | [0.408, 0.939] |
| Prospective | cg11248361 | DMPs | FEV-1/FVC ratio 2001 | 1-SD higher methylation beta | Beta | 0.008 | 0.004 | 3.3971E-02 | [0.001, 0.015] |
| Prospective | cg11795976 | DMPs | FEV-1 % predicted 2016 | 1-SD higher methylation beta | Beta | 3.906 | 1.537 | 1.1528E-02 | [0.893, 6.920] |
| Prospective | cg11795976 | DMPs | Byssinosis 2001 | 1-SD higher methylation beta | OR | 3.249 |  | 1.5412E-02 | [1.252, 8.427] |
| Prospective | cg11795976 | DMPs | FVC % predicted 2016 | 1-SD higher methylation beta | Beta | 3.287 | 1.459 | 2.4883E-02 | [0.429, 6.146] |
| Prospective | cg12318285 | DMPs | FEV-1/FVC < GLI LLN 2006 | 1-SD higher methylation beta | OR | 1.791 |  | 3.5171E-02 | [1.041, 3.082] |
| Prospective | cg12318285 | DMPs | FVC (mL) 2001 | 1-SD higher methylation beta | Beta | -50.163 | 24.525 | 4.1331E-02 | [-98.232, -2.095] |
| Prospective | cg15499393 | DMPs | FEV-1/FVC ratio 2016 | 1-SD higher methylation beta | Beta | 0.012 | 0.006 | 4.7981E-02 | [0.000, 0.023] |
| Prospective | cg16290931 | DMPs | Annual FEV-1 change 1996–2001 | 1-SD higher methylation beta | Beta | 4.464 | 1.902 | 1.9363E-02 | [0.736, 8.193] |
| Prospective | cg16371865 | DMPs | FEV-1/FVC ratio 2016 | 1-SD higher methylation beta | Beta | 0.009 | 0.004 | 1.8192E-02 | [0.002, 0.016] |
| Prospective | cg16371865 | DMPs | FEV-1 (mL) 2016 | 1-SD higher methylation beta | Beta | 52.024 | 22.740 | 2.2807E-02 | [7.454, 96.594] |
| Prospective | cg16371865 | DMPs | FEV-1 % predicted 2016 | 1-SD higher methylation beta | Beta | 2.019 | 0.950 | 3.4329E-02 | [0.157, 3.881] |
| Prospective | cg19055984 | DMPs | Chronic bronchitis 2001 | 1-SD higher methylation beta | OR | 0.643 |  | 2.5126E-02 | [0.436, 0.946] |
| Prospective | cg19055984 | DMPs | FEV-1/FVC ratio 2006 | 1-SD higher methylation beta | Beta | -0.009 | 0.004 | 2.6069E-02 | [-0.016, -0.001] |
| Prospective | cg19055984 | DMPs | FEV-1 % predicted 2006 | 1-SD higher methylation beta | Beta | -1.637 | 0.746 | 2.8873E-02 | [-3.100, -0.174] |
| Prospective | cg19055984 | DMPs | FEV-1/FVC < GLI LLN 2006 | 1-SD higher methylation beta | OR | 1.580 |  | 4.5806E-02 | [1.009, 2.475] |
| Prospective | cg19925755 | DMPs | Annual FVC change 1996–2006 | 1-SD higher methylation beta | Beta | -3.348 | 1.509 | 2.7056E-02 | [-6.305, -0.391] |
| Prospective | cg22740441 | DMPs | FEV-1 % predicted 2016 | 1-SD higher methylation beta | Beta | 3.033 | 1.528 | 4.8060E-02 | [0.037, 6.029] |
| Prospective | cg22766862 | DMPs | Annual FVC change 1996–2011 | 1-SD higher methylation beta | Beta | -2.505 | 1.157 | 3.1029E-02 | [-4.774, -0.237] |
| Prospective | cg25022932 | DMPs | FEV-1/FVC ratio 2001 | 1-SD higher methylation beta | Beta | 0.007 | 0.003 | 3.1120E-02 | [0.001, 0.014] |
| Prospective | cg26277709 | DMPs | FEV-1/FVC ratio 2001 | 1-SD higher methylation beta | Beta | 0.007 | 0.003 | 2.7240E-02 | [0.001, 0.014] |
| Prospective | cg26277709 | DMPs | FEV-1/FVC <0.70 2011 | 1-SD higher methylation beta | OR | 0.669 |  | 3.5456E-02 | [0.460, 0.973] |
| Prospective | cg26491213 | DMPs | Chronic cough 2001 | 1-SD higher methylation beta | OR | 0.607 |  | 1.3478E-02 | [0.408, 0.902] |
| Prospective | cg26491213 | DMPs | FEV-1/FVC <0.70 2006 | 1-SD higher methylation beta | OR | 0.677 |  | 2.6005E-02 | [0.480, 0.954] |
| Prospective | cg26491213 | DMPs | FEV-1/FVC < GLI LLN 2001 | 1-SD higher methylation beta | OR | 0.625 |  | 3.8361E-02 | [0.400, 0.975] |
| Prospective | cg26491213 | DMPs | Chronic bronchitis 2001 | 1-SD higher methylation beta | OR | 0.667 |  | 4.0354E-02 | [0.453, 0.982] |
| Prospective | cg26491213 | DMPs | FEV-1 % predicted 2006 | 1-SD higher methylation beta | Beta | 1.468 | 0.721 | 4.2494E-02 | [0.054, 2.882] |
| Prospective | cg26491213 | DMPs | FVC % predicted 2006 | 1-SD higher methylation beta | Beta | 1.278 | 0.648 | 4.9237E-02 | [0.008, 2.548] |
| Prospective | IRS<sub>(Hillary-BayesPR)</sub> | IRS | FEV-1 (mL) 2001 | 1-SD higher score | Beta | -56.582 | 16.994 | 9.3377E-04 | [-89.890, -23.274] |
| Prospective | IRS<sub>(Hillary-BayesPR)</sub> | IRS | FEV-1 % predicted 2001 | 1-SD higher score | Beta | -1.952 | 0.621 | 1.7591E-03 | [-3.169, -0.735] |
| Prospective | IRS<sub>(Hillary-BayesPR)</sub> | IRS | FVC (mL) 2016 | 1-SD higher score | Beta | -76.760 | 27.934 | 6.3421E-03 | [-131.511, -22.008] |
| Prospective | IRS<sub>(Hillary-BayesPR)</sub> | IRS | Dyspnea 2001 | 1-SD higher score | OR | 1.440 |  | 6.4571E-03 | [1.108, 1.872] |
| Prospective | IRS<sub>(Hillary-BayesPR)</sub> | IRS | FVC (mL) 2001 | 1-SD higher score | Beta | -51.857 | 19.388 | 7.7215E-03 | [-89.857, -13.857] |
| Prospective | IRS<sub>(Hillary-BayesPR)</sub> | IRS | FEV-1 % predicted 2011 | 1-SD higher score | Beta | -2.034 | 0.796 | 1.0958E-02 | [-3.593, -0.474] |
| Prospective | IRS<sub>(Hillary-BayesPR)</sub> | IRS | FEV-1 (mL) 2011 | 1-SD higher score | Beta | -51.360 | 20.451 | 1.2426E-02 | [-91.444, -11.277] |
| Prospective | IRS<sub>(Hillary-BayesPR)</sub> | IRS | FVC (mL) 2011 | 1-SD higher score | Beta | -60.724 | 24.307 | 1.2890E-02 | [-108.367, -13.082] |
| Prospective | IRS<sub>(Hillary-BayesPR)</sub> | IRS | FVC % predicted 2001 | 1-SD higher score | Beta | -1.382 | 0.555 | 1.3060E-02 | [-2.470, -0.295] |
| Prospective | IRS<sub>(Hillary-BayesPR)</sub> | IRS | FEV-1 (mL) 2016 | 1-SD higher score | Beta | -56.910 | 23.216 | 1.4773E-02 | [-102.415, -11.406] |
| Prospective | IRS<sub>(Hillary-BayesPR)</sub> | IRS | FEV-1/FVC < GLI LLN 2001 | 1-SD higher score | OR | 1.654 |  | 1.4804E-02 | [1.104, 2.480] |
| Prospective | IRS<sub>(Hillary-BayesPR)</sub> | IRS | FEV-1 (mL) 2006 | 1-SD higher score | Beta | -47.408 | 19.442 | 1.5166E-02 | [-85.515, -9.301] |
| Prospective | IRS<sub>(Hillary-BayesPR)</sub> | IRS | FVC % predicted 2016 | 1-SD higher score | Beta | -2.251 | 0.926 | 1.5647E-02 | [-4.066, -0.435] |
| Prospective | IRS<sub>(Hillary-BayesPR)</sub> | IRS | FVC % predicted 2011 | 1-SD higher score | Beta | -1.742 | 0.725 | 1.6715E-02 | [-3.163, -0.321] |
| Prospective | IRS<sub>(Hillary-BayesPR)</sub> | IRS | FEV-1 % predicted 2006 | 1-SD higher score | Beta | -1.659 | 0.715 | 2.0870E-02 | [-3.061, -0.257] |
| Prospective | IRS<sub>(Hillary-BayesPR)</sub> | IRS | FEV-1/FVC < GLI LLN 2006 | 1-SD higher score | OR | 1.656 |  | 2.1448E-02 | [1.077, 2.544] |
| Prospective | IRS<sub>(Hillary-BayesPR)</sub> | IRS | FEV-1/FVC <0.70 2016 | 1-SD higher score | OR | 1.713 |  | 2.2921E-02 | [1.077, 2.725] |
| Prospective | IRS<sub>(Hillary-BayesPR)</sub> | IRS | FEV-1 % predicted 2016 | 1-SD higher score | Beta | -2.224 | 0.980 | 2.3871E-02 | [-4.144, -0.304] |
| Prospective | IRS<sub>(Hillary-BayesPR)</sub> | IRS | Chronic bronchitis 2001 | 1-SD higher score | OR | 1.486 |  | 2.6844E-02 | [1.046, 2.109] |
| Prospective | IRS<sub>(Hillary-BayesPR)</sub> | IRS | FEV-1/FVC <0.70 2001 | 1-SD higher score | OR | 1.451 |  | 2.9322E-02 | [1.038, 2.028] |
| Prospective | IRS<sub>(Hillary-elastic-net)</sub> | IRS | FVC (mL) 2016 | 1-SD higher score | Beta | -96.097 | 28.682 | 9.0444E-04 | [-152.314, -39.880] |
| Prospective | IRS<sub>(Hillary-elastic-net)</sub> | IRS | FEV-1 (mL) 2001 | 1-SD higher score | Beta | -54.698 | 17.431 | 1.8008E-03 | [-88.863, -20.534] |
| Prospective | IRS<sub>(Hillary-elastic-net)</sub> | IRS | FEV-1 (mL) 2016 | 1-SD higher score | Beta | -74.656 | 23.833 | 1.8948E-03 | [-121.368, -27.944] |
| Prospective | IRS<sub>(Hillary-elastic-net)</sub> | IRS | FVC % predicted 2016 | 1-SD higher score | Beta | -2.873 | 0.951 | 2.7320E-03 | [-4.738, -1.008] |
| Prospective | IRS<sub>(Hillary-elastic-net)</sub> | IRS | FEV-1 % predicted 2001 | 1-SD higher score | Beta | -1.904 | 0.636 | 2.9011E-03 | [-3.151, -0.657] |
| Prospective | IRS<sub>(Hillary-elastic-net)</sub> | IRS | FEV-1 % predicted 2016 | 1-SD higher score | Beta | -2.904 | 1.006 | 4.1613E-03 | [-4.877, -0.932] |
| Prospective | IRS<sub>(Hillary-elastic-net)</sub> | IRS | FEV-1 (mL) 2011 | 1-SD higher score | Beta | -56.211 | 20.786 | 7.1440E-03 | [-96.952, -15.470] |
| Prospective | IRS<sub>(Hillary-elastic-net)</sub> | IRS | FEV-1 % predicted 2011 | 1-SD higher score | Beta | -2.169 | 0.805 | 7.3738E-03 | [-3.748, -0.591] |
| Prospective | IRS<sub>(Hillary-elastic-net)</sub> | IRS | FVC (mL) 2011 | 1-SD higher score | Beta | -63.894 | 24.724 | 1.0117E-02 | [-112.352, -15.436] |
| Prospective | IRS<sub>(Hillary-elastic-net)</sub> | IRS | Dyspnea 2001 | 1-SD higher score | OR | 1.415 |  | 1.0641E-02 | [1.084, 1.848] |
| Prospective | IRS<sub>(Hillary-elastic-net)</sub> | IRS | FEV-1 (mL) 2006 | 1-SD higher score | Beta | -49.754 | 19.962 | 1.3069E-02 | [-88.879, -10.629] |
| Prospective | IRS<sub>(Hillary-elastic-net)</sub> | IRS | FVC % predicted 2011 | 1-SD higher score | Beta | -1.820 | 0.734 | 1.3563E-02 | [-3.259, -0.382] |
| Prospective | IRS<sub>(Hillary-elastic-net)</sub> | IRS | FVC (mL) 2001 | 1-SD higher score | Beta | -49.144 | 19.876 | 1.3743E-02 | [-88.101, -10.188] |
| Prospective | IRS<sub>(Hillary-elastic-net)</sub> | IRS | FEV-1/FVC <0.70 2001 | 1-SD higher score | OR | 1.527 |  | 1.7140E-02 | [1.078, 2.163] |
| Prospective | IRS<sub>(Hillary-elastic-net)</sub> | IRS | FVC % predicted 2001 | 1-SD higher score | Beta | -1.350 | 0.568 | 1.7916E-02 | [-2.464, -0.236] |
| Prospective | IRS<sub>(Hillary-elastic-net)</sub> | IRS | FEV-1 % predicted 2006 | 1-SD higher score | Beta | -1.709 | 0.732 | 2.0124E-02 | [-3.144, -0.273] |
| Prospective | IRS<sub>(Hillary-elastic-net)</sub> | IRS | FEV-1/FVC <0.70 2016 | 1-SD higher score | OR | 1.748 |  | 2.0155E-02 | [1.091, 2.800] |
| Prospective | IRS<sub>(Stevenson)</sub> | IRS | FEV-1/FVC <0.70 2016 | 1-SD higher score | OR | 2.595 |  | 3.9690E-03 | [1.356, 4.965] |
| Prospective | IRS<sub>(Stevenson)</sub> | IRS | FEV-1/FVC < GLI LLN 2011 | 1-SD higher score | OR | 2.689 |  | 1.0943E-02 | [1.255, 5.759] |
| Prospective | IRS<sub>(Stevenson)</sub> | IRS | FEV-1/FVC <0.70 2011 | 1-SD higher score | OR | 1.983 |  | 1.2879E-02 | [1.156, 3.402] |
| Prospective | IRS<sub>(Stevenson)</sub> | IRS | FEV-1/FVC ratio 2011 | 1-SD higher score | Beta | -0.013 | 0.005 | 1.6166E-02 | [-0.023, -0.002] |
| Prospective | IRS<sub>(Stevenson)</sub> | IRS | FEV-1/FVC < GLI LLN 2016 | 1-SD higher score | OR | 2.451 |  | 3.4900E-02 | [1.066, 5.639] |
| Prospective | IRS<sub>(Stevenson)</sub> | IRS | Annual FVC change 1996–2001 | 1-SD higher score | Beta | -7.901 | 3.777 | 3.6970E-02 | [-15.303, -0.499] |
| Prospective | IRS<sub>(Ligthart)</sub> | IRS | FEV-1 (mL) 2016 | 1-SD higher score | Beta | -68.726 | 25.781 | 8.0753E-03 | [-119.258, -18.195] |
| Prospective | IRS<sub>(Ligthart)</sub> | IRS | FEV-1 % predicted 2016 | 1-SD higher score | Beta | -2.745 | 1.074 | 1.1026E-02 | [-4.849, -0.641] |
| Prospective | IRS<sub>(Ligthart)</sub> | IRS | Annual FVC change 1996–2016 | 1-SD higher score | Beta | -2.402 | 1.003 | 1.7257E-02 | [-4.368, -0.436] |
| Prospective | IRS<sub>(Ligthart)</sub> | IRS | FVC (mL) 2016 | 1-SD higher score | Beta | -73.203 | 31.171 | 1.9467E-02 | [-134.299, -12.108] |
| Prospective | IRS<sub>(Ligthart)</sub> | IRS | FVC % predicted 2016 | 1-SD higher score | Beta | -2.328 | 1.018 | 2.2911E-02 | [-4.324, -0.332] |
| Prospective | IRS<sub>(Ligthart)</sub> | IRS | Annual FVC change 1996–2001 | 1-SD higher score | Beta | -6.429 | 2.851 | 2.4578E-02 | [-12.016, -0.842] |
| Prospective | IRS<sub>(Ligthart)</sub> | IRS | FVC % predicted 2001 | 1-SD higher score | Beta | -1.389 | 0.625 | 2.6579E-02 | [-2.614, -0.165] |
| Prospective | IRS<sub>(Ligthart)</sub> | IRS | FEV-1 % predicted 2001 | 1-SD higher score | Beta | -1.491 | 0.706 | 3.5074E-02 | [-2.874, -0.108] |
| Prospective | IRS<sub>(Ligthart)</sub> | IRS | Annual FEV-1 change 1996–2016 | 1-SD higher score | Beta | -1.473 | 0.725 | 4.3112E-02 | [-2.895, -0.052] |
| Prospective | IRS<sub>(Ligthart)</sub> | IRS | FEV-1/FVC <0.70 2016 | 1-SD higher score | OR | 1.571 |  | 4.5960E-02 | [1.008, 2.449] |
| Prospective | IRS<sub>(Wielscher)</sub> | IRS | FEV-1 % predicted 2016 | 1-SD higher score | Beta | -4.190 | 1.372 | 2.4531E-03 | [-6.880, -1.500] |
| Prospective | IRS<sub>(Wielscher)</sub> | IRS | FEV-1/FVC <0.70 2016 | 1-SD higher score | OR | 2.156 |  | 7.7800E-03 | [1.224, 3.797] |
| Prospective | IRS<sub>(Wielscher)</sub> | IRS | FEV-1 (mL) 2016 | 1-SD higher score | Beta | -87.994 | 33.158 | 8.3611E-03 | [-152.984, -23.004] |
| Prospective | IRS<sub>(Wielscher)</sub> | IRS | FVC % predicted 2016 | 1-SD higher score | Beta | -3.428 | 1.304 | 8.9714E-03 | [-5.984, -0.873] |
| Prospective | IRS<sub>(Wielscher)</sub> | IRS | FEV-1 % predicted 2011 | 1-SD higher score | Beta | -2.844 | 1.142 | 1.3146E-02 | [-5.082, -0.606] |
| Prospective | IRS<sub>(Wielscher)</sub> | IRS | FEV-1 % predicted 2001 | 1-SD higher score | Beta | -2.193 | 0.924 | 1.7987E-02 | [-4.004, -0.382] |
| Prospective | IRS<sub>(Wielscher)</sub> | IRS | FVC (mL) 2016 | 1-SD higher score | Beta | -94.495 | 40.084 | 1.9008E-02 | [-173.059, -15.932] |
| Prospective | IRS<sub>(Wielscher)</sub> | IRS | FVC % predicted 2001 | 1-SD higher score | Beta | -1.831 | 0.822 | 2.6433E-02 | [-3.442, -0.219] |
| Prospective | IRS<sub>(Wielscher)</sub> | IRS | FEV-1 (mL) 2011 | 1-SD higher score | Beta | -59.885 | 29.926 | 4.6070E-02 | [-118.540, -1.230] |

Separate multivariable linear regression models for continuous outcomes and logistic regression models for binary outcomes were adjusted for log cumulative endotoxin, baseline age, sex, height, years since cessation of endotoxin exposure, smoking pack-years, body mass index, EpiDISH-estimated immune-cell fractions, and Sample Plate. Age, sex, and height were omitted from models of GLI percent-predicted FEV-1 and FVC because these characteristics are incorporated in the reference equations. DNAm biomarkers and covariates were measured in 1996; concurrent outcomes were measured in 1996 and prospective outcomes in 2001, 2006, 2011, or 2016. DMP associations are expressed per 1-SD higher methylation beta value and IRS associations per 1-SD higher score. Continuous-outcome effects are beta coefficients; binary-outcome effects are odds ratios. The table includes nominal associations at P<0.05 with 95% model-based confidence intervals. Abbreviations: BMI=body mass index; DMP=differentially methylated position; DNAm=DNA methylation; FEV-1=forced expiratory volume in 1 second; FVC=forced vital capacity; GLI=Global Lung Function Initiative; IRS=inflammation risk score; SD=standard deviation.

**Supplemental Figure 1.**
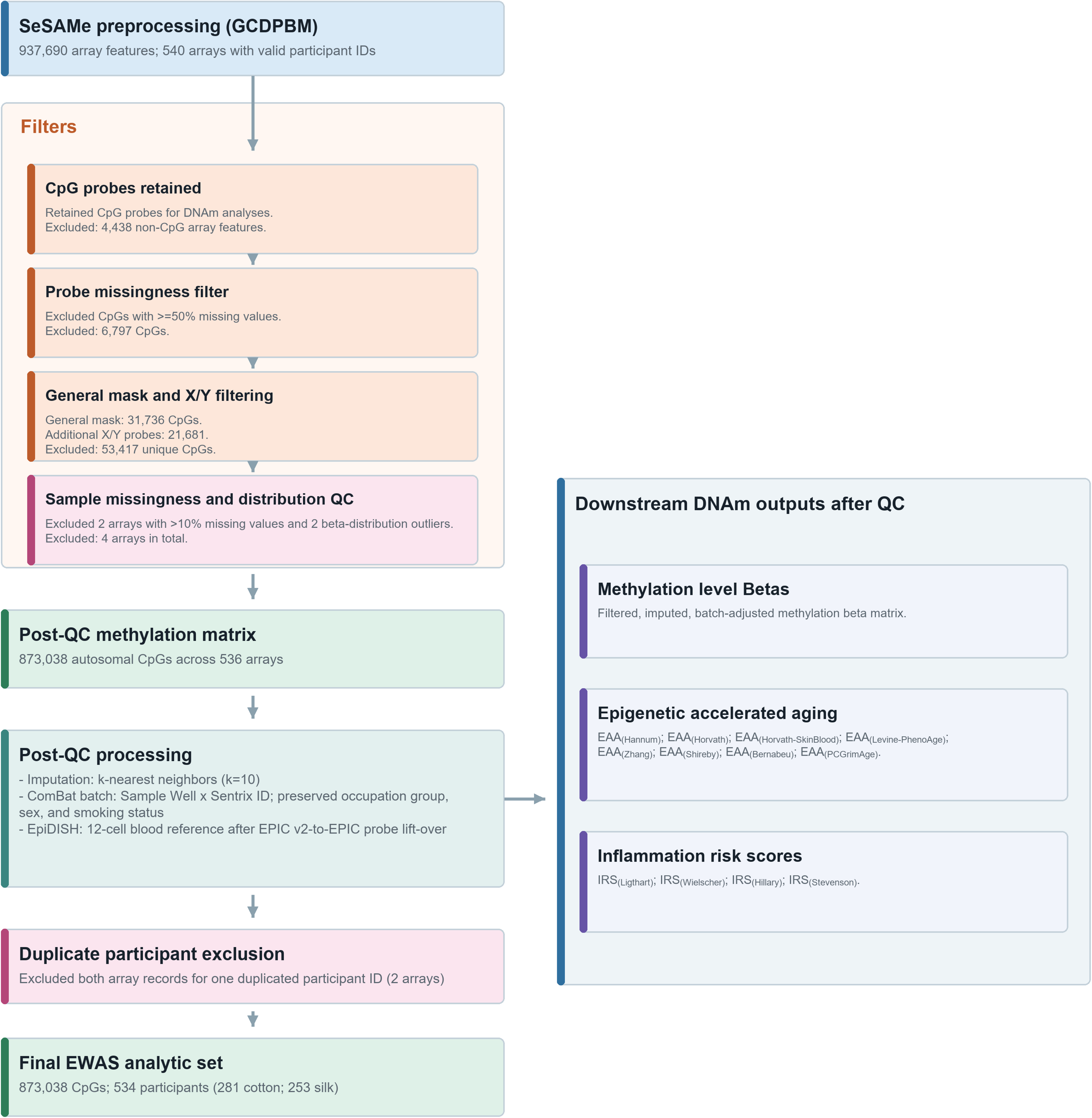
DNA methylation data quality-control workflow. Downstream DNAm outputs after QC Methylation level Betas. Filtered, imputed, batch-adjusted methylation beta matrix. **Epigenetic accelerated aging** EAA(Hannum); EAA(Horvath); EAA(Horvath-SkinBlood); EAA(Levine-PhenoAge); EAA(Zhang); EAA(Shireby); EAA(Bernabeu); EAA(PCGrimAge)-**Inflammation risk scores** I RS(Ligthart); IRS(wielscher); I RS(Hillary); I RS(stevenson)-Raw IDATs were processed in SeSAMe using the GCDPBM preprocessing sequence. The STWS EPIC v2 matrix included 937,690 array features across 540 arrays linked to valid participant IDs. Probe filtering removed 4,438 non-CpG features and 6,797 CpGs with >=50% missing values. The EPIC v2 general mask removed 31,736 CpGs, followed by 21,681 additional X/Y probes (53,417 unique exclusions), leaving 873,038 autosomal CpGs. Sample QC removed two arrays with >10% missing values and two beta-distribution outliers, leaving 536 post-QC arrays. Both records for one duplicated participant ID were excluded before the EWAS, leaving 534 participants (281 cotton; 253 silk). Missing values were imputed by k-nearest neighbors (k=10). ComBat used Sample Well x Sentrix ID as the batch variable while preserving occupation group, sex, and smoking status. EpiDISH used a 12-cell blood reference after EPIC v2-to-EPIC probe lift-over. Abbreviations: ComBat=batch-effect correction using empirical Bayes; DNAm=DNA methylation; EAA=epigenetic age acceleration; EpiDISH=Epigenetic Dissection of Intra-Sample Heterogeneity; IRS=inflammation risk score; QC=aualitv control: STWS=Shanahai Textile Worker Study.

**Supplemental Figure 2.**
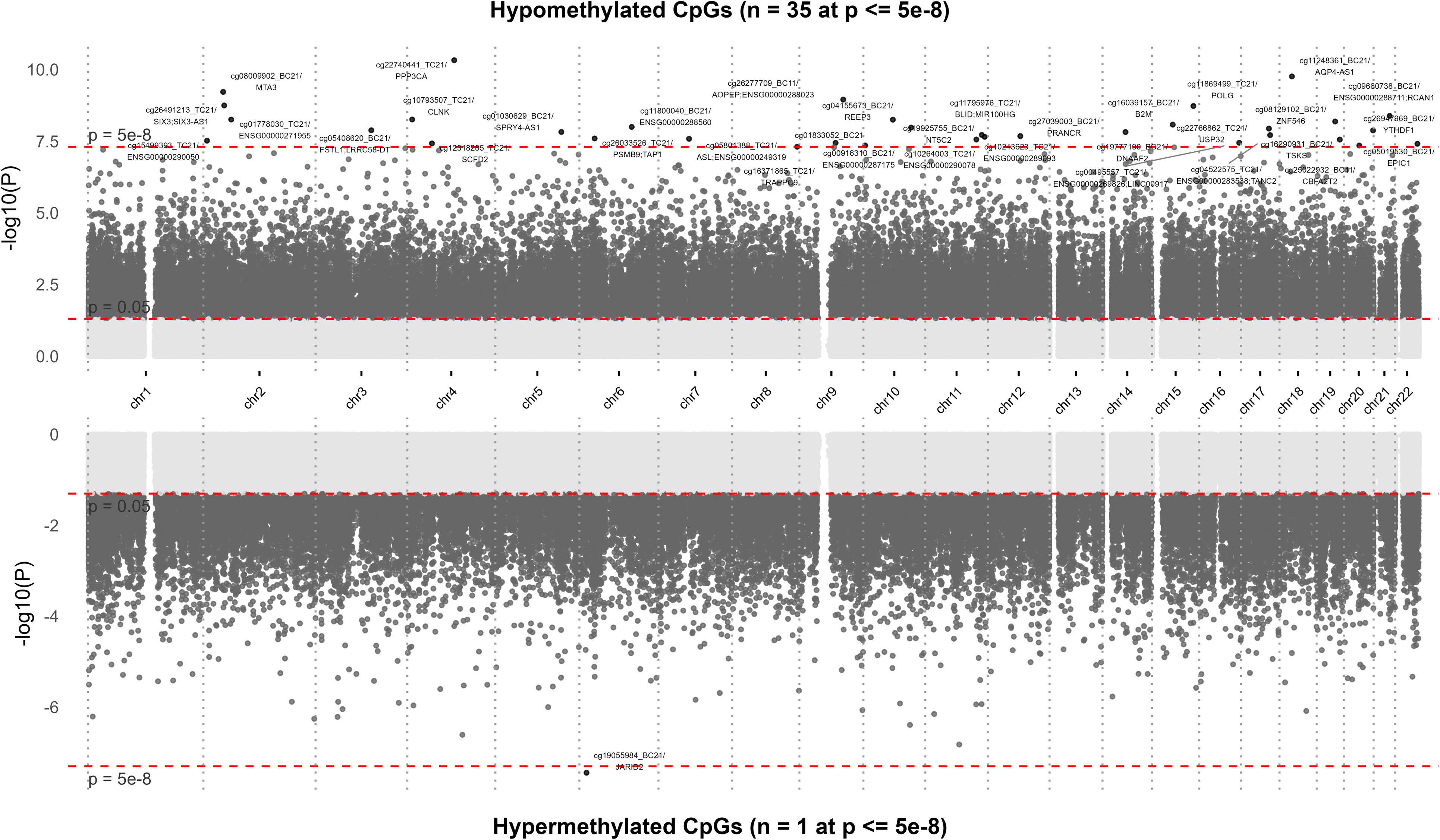
Miami plot of statistical significance for endotoxin-related cytosine-phosphate-guanine loci stratified by hypomethylated and hypermethylated associations. Abbreviations: CpG=cytosine-phosphate-guanine.

**Supplemental Figure 3.**
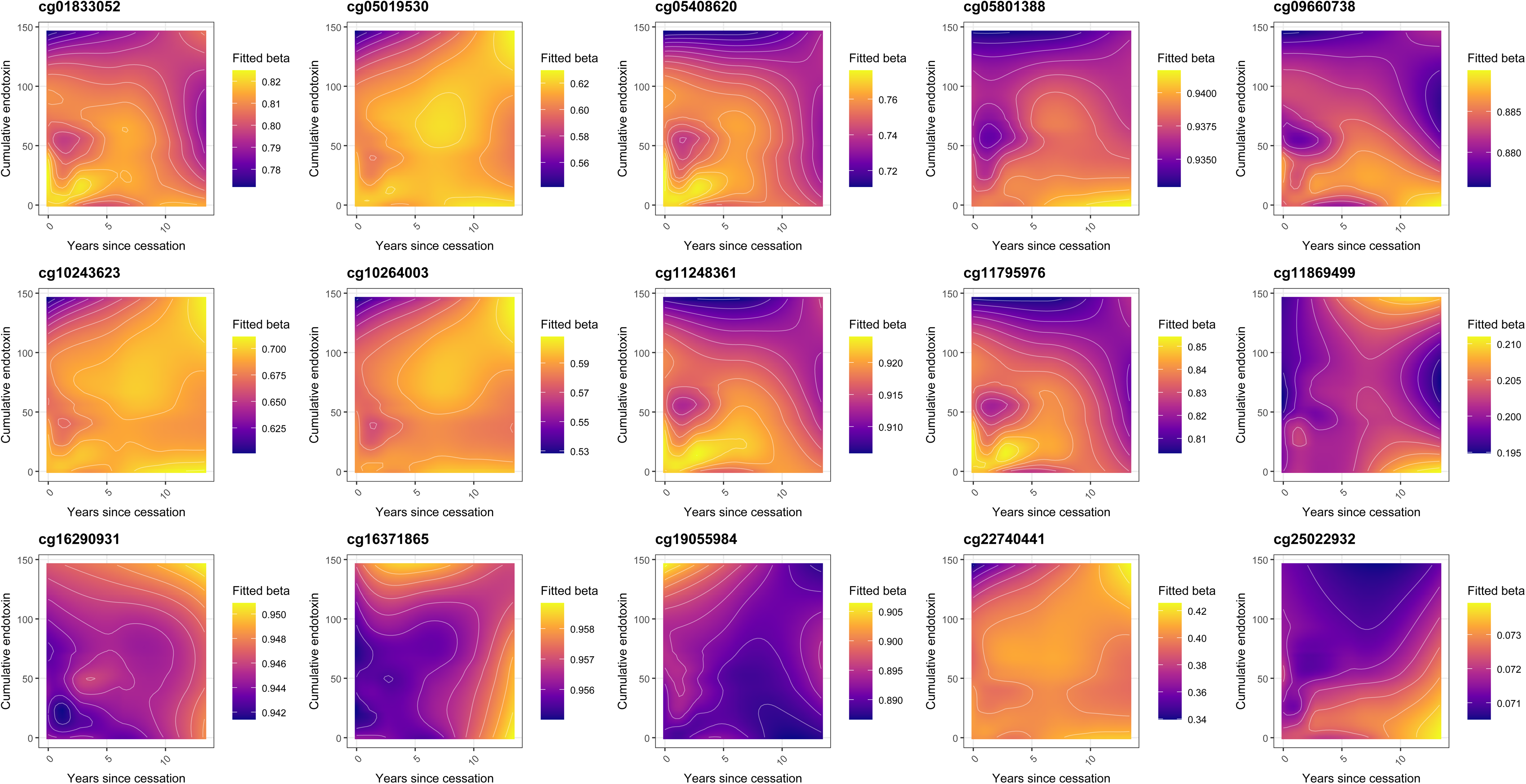
Contour plots of 15 differentially methylated positions associated with endotoxin that also showed significant associations with years since cessation from endotoxin exposure. Cumulative endotoxin is shown on the y-axis, years since cessation are shown on the x-axis, and color represents adjusted methylation beta values. Abbreviations: DMP=differentially methylated position.

**Supplemental Figure 4.**
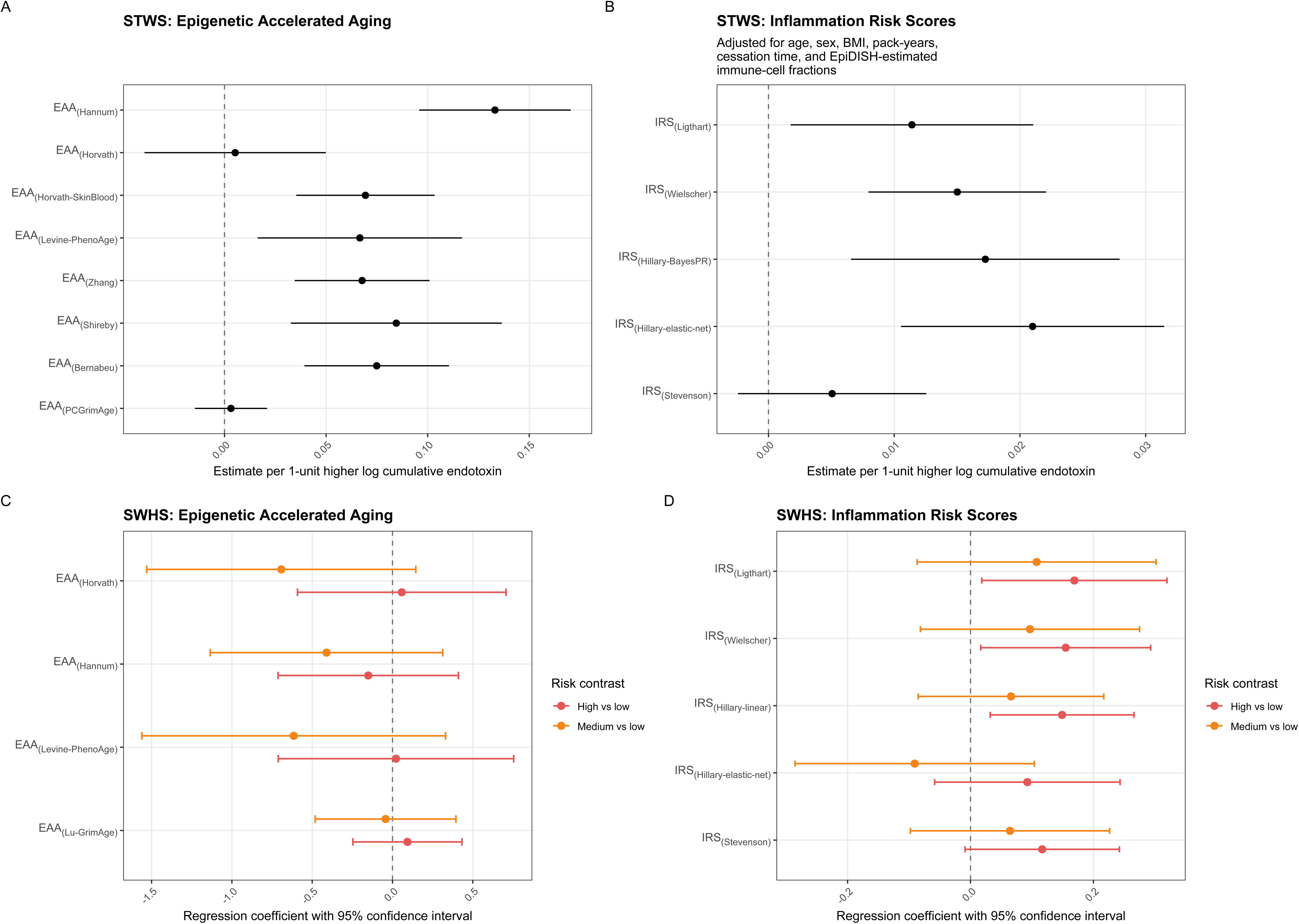
Endotoxin associations with inflammation risk scores and epigenetic accelerated aging. A) In STWS, epigenetic age acceleration was defined as the residual from regression of each epigenetic age measure on chronological age; residuals were modeled with log cumulative endotoxin, age, sex, body mass index, smoking pack-years, years since exposure cessation, and EpiDISH-estimated immune-cell fractions. B) In STWS, standardized inflammation risk scores were modeled with log cumulative endotoxin and the same covariates. C) In SWHS, epigenetic age acceleration was modeled with occupational endotoxin-risk category, age, body mass index, education, case-control status, EpiDISH-estimated immune-cell fractions, and Sample Well, with a random intercept for MatchSet. D) In SWHS, inflammation risk scores were modeled with occupational endotoxin-risk category using the same covariates and random intercept as in panel C. IRS_(Stevenson)_ is an interleukin-6-based score; the other inflammation risk scores are C-reactive protein-based scores. Abbreviations: BayesPR=Bayesian penalized regression; EAA=epigenetic age acceleration; EpiDISH=Epigenetic Dissection of Intra-Sample Heterogeneity; IRS=inflammation risk score; STWS=Shanghai Textile Worker Study; SWHS=Shanghai Women’s Health Study.

**Supplemental Figure 5.**
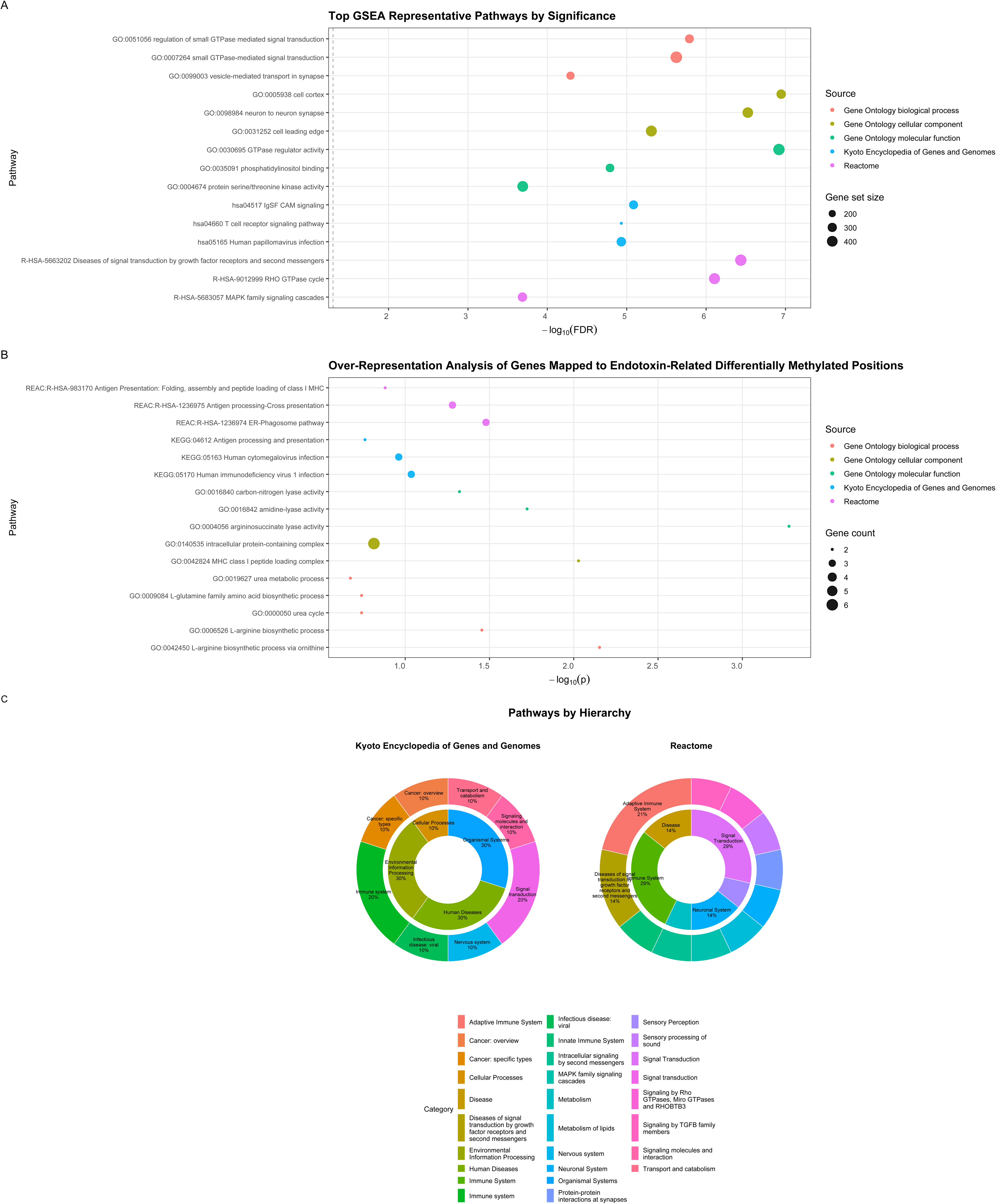
Pathway enrichment results for endotoxin-related DNA methylation. A) The three most significant collapsed representative pathways from each Gene Ontology domain, Kyoto Encyclopedia of Genes and Genomes, and Reactome are shown; the x-axis gives -log10(false discovery rate), point size indicates gene-set size, and color indicates source. For GSEA, CpGs were inverse-variance pooled by gene and ranked by unsigned -log10(P). B) Over-Representation Analysis of genes mapped to the 36 differentially methylated positions. C) Broad distribution of Kyoto Encyclopedia of Genes and Genomes and Reactome pathways across two hierarchy levels after pathway collapsing. Abbreviations: CpG=cytosine-phosphate-guanine; DMP=differentially methylated position; FDR=false discovery rate; GO=Gene Ontology; GSEA=Gene Set Enrichment Analysis; KEGG=Kyoto Encyclopedia of Genes and Genomes; ORA=Over-Representation Analysis.

| Occupation code | Occupation | Ordinal exposure-intensity rank | Broad group | Broad-group ordinal exposure-intensity rank |
| --- | --- | --- | --- | --- |
| S_BL_OCC_3D_EN079 | Ever/Never Veterinarians | High | Professional / technical / medical / education / arts | Low |
| S_BL_OCC_3D_EN433 | Ever/Never Recovery of Waste Materials | High | Waste material recovery | High |
| S_BL_OCC_3D_EN522 | Ever/Never Cleaning/Waste Disposal | High | Cleaning / waste disposal | High |
| S_BL_OCC_3D_EN600 | Ever/Never Agric/Forestry/Animal Husbandry | High | Agriculture / forestry / animal husbandry (broad header) | High |
| S_BL_OCC_3D_EN611 | Ever/Never Food Crops Farm Workers | High | Crop agriculture | High |
| S_BL_OCC_3D_EN612 | Ever/Never Cotton Plantation Workers | High | Crop agriculture | High |
| S_BL_OCC_3D_EN613 | Ever/Never Vegetables Farm Workers | High | Crop agriculture | High |
| S_BL_OCC_3D_EN614 | Ever/Never Tea Plantation/Orchard Farmers | High | Crop agriculture | High |
| S_BL_OCC_3D_EN619 | Ever/Never Other Farm Workers | High | Crop agriculture | High |
| S_BL_OCC_3D_EN631 | Ever/Never Large Livestock Raisers | High | Livestock / animal husbandry | High |
| S_BL_OCC_3D_EN632 | Ever/Never Domesticated Birds/Animals Raisers | High | Livestock / animal husbandry | High |
| S_BL_OCC_3D_EN633 | Ever/Never Raisers Special-purposes Animals | High | Livestock / animal husbandry | High |
| S_BL_OCC_3D_EN634 | Ever/Never Dvlpmnt Animal Specs/Body Process | High | Livestock / animal husbandry | High |
| S_BL_OCC_3D_EN639 | Ever/Never Other Animal Husbandry Workers | High | Livestock / animal husbandry | High |
| S_BL_OCC_3D_EN661 | Ever/Never Operators of Tractors | High | Farm machinery / field operations | Medium |
| S_BL_OCC_3D_EN669 | Ever/Never Other Operators of Farm Machinery | High | Farm machinery / field operations | Medium |
| S_BL_OCC_3D_EN699 | Ever/Never Other Agric/Forstry/Animl Husbandry | High | Other agriculture / forestry / animal husbandry | Medium |
| S_BL_OCC_3D_EN751 | Ever/Never Fibers Pre-Treatment Workers | High | Textile / fiber processing | High |
| S_BL_OCC_3D_EN752 | Ever/Never Spinning/Filature/Plying/Twisting | High | Textile / fiber processing | High |
| S_BL_OCC_3D_EN753 | Ever/Never Instal/Maint Mach Weavng/Knitng | High | Textile / fiber processing | High |
| S_BL_OCC_3D_EN754 | Ever/Never Cloth-Weaving Workers | High | Textile / fiber processing | High |
| S_BL_OCC_3D_EN755 | Ever/Never Knitting Workers | High | Textile / fiber processing | High |
| S_BL_OCC_3D_EN756 | Ever/Never Textile Printing & Dyeing Workers | High | Textile / fiber processing | High |
| S_BL_OCC_3D_EN759 | Ever/Never Other Spinng/Weavng/Knitng/Printng | High | Textile / fiber processing | High |
| S_BL_OCC_3D_EN761 | Ever/Never Production of leather/fur | High | Leather / fur / feather processing | Medium |
| S_BL_OCC_3D_EN764 | Ever/Never Pre-Processing Feather & Stuffing | High | Leather / fur / feather processing | Medium |
| S_BL_OCC_3D_EN781 | Ever/Never Rice/Flour Millers & Oil Pressers | High | Raw food / grain / animal-food processing | High |
| S_BL_OCC_3D_EN783 | Ever/Never Butchers, Animal-Food Processers | High | Raw food / grain / animal-food processing | High |
| S_BL_OCC_3D_EN787 | Ever/Never Tea Processors | High | Other food / beverage / tobacco processing | Medium |
| S_BL_OCC_3D_EN790 | Ever/Never Tobacco Production Workers | High | Other food / beverage / tobacco processing | Medium |
| S_BL_OCC_3D_EN801 | Ever/Never Timber Pretreatment Workers | High | Wood / paper processing | Medium |
| S_BL_OCC_3D_EN805 | Ever/Never Fin Wood,Bamboo,Jute,Rattan,Palm | High | Wood / paper processing | Medium |
| S_BL_OCC_3D_EN011 | Ever/Never Researchers in Philosophy | Low | Professional / technical / medical / education / arts | Low |
| S_BL_OCC_3D_EN012 | Ever/Never Researchers in Sociology | Low | Professional / technical / medical / education / arts | Low |
| S_BL_OCC_3D_EN013 | Ever/Never Researchers in Economics | Low | Professional / technical / medical / education / arts | Low |
| S_BL_OCC_3D_EN014 | Ever/Never Researchers in Law | Low | Professional / technical / medical / education / arts | Low |
| S_BL_OCC_3D_EN015 | Ever/Never Researchers in Education | Low | Professional / technical / medical / education / arts | Low |
| S_BL_OCC_3D_EN016 | Ever/Never Researchers in Literature & Arts | Low | Professional / technical / medical / education / arts | Low |
| S_BL_OCC_3D_EN017 | Ever/Never Researchers Library Sci/Info Tech | Low | Professional / technical / medical / education / arts | Low |
| S_BL_OCC_3D_EN018 | Ever/Never Researchers in History | Low | Professional / technical / medical / education / arts | Low |
| S_BL_OCC_3D_EN020 | Ever/Never Researchers in Engineering | Low | Professional / technical / medical / education / arts | Low |
| S_BL_OCC_3D_EN029 | Ever/Never Researchers in Other Sciences | Low | Professional / technical / medical / education / arts | Low |
| S_BL_OCC_3D_EN031 | Ever/Never Urban Develop & Planning Persnl | Low | Professional / technical / medical / education / arts | Low |
| S_BL_OCC_3D_EN032 | Ever/Never Civil Engineering Tech Personnel | Low | Professional / technical / medical / education / arts | Low |
| S_BL_OCC_3D_EN033 | Ever/Never Electrical/Electronic Engineering | Low | Professional / technical / medical / education / arts | Low |
| S_BL_OCC_3D_EN034 | Ever/Never Mechanical Engineering Personnel | Low | Professional / technical / medical / education / arts | Low |
| S_BL_OCC_3D_EN035 | Ever/Never Chemical Engineering Personnel | Low | Professional / technical / medical / education / arts | Low |
| S_BL_OCC_3D_EN036 | Ever/Never Metallurgical Engin Personnel | Low | Professional / technical / medical / education / arts | Low |
| S_BL_OCC_3D_EN037 | Ever/Never Mining Engineering Personnel | Low | Professional / technical / medical / education / arts | Low |
| S_BL_OCC_3D_EN038 | Ever/Never Indust Management Tech Personnel | Low | Professional / technical / medical / education / arts | Low |
| S_BL_OCC_3D_EN039 | Ever/Never Mapping/Hydrology Tech Personnel | Low | Professional / technical / medical / education / arts | Low |
| S_BL_OCC_3D_EN040 | Ever/Never Meteorology/Earthquake Tech Persnl | Low | Professional / technical / medical / education / arts | Low |
| S_BL_OCC_3D_EN041 | Ever/Never Geological Engineering Tech Persnl | Low | Professional / technical / medical / education / arts | Low |
| S_BL_OCC_3D_EN045 | Ever/Never Internet/Computer Hardware/Software | Low | Professional / technical / medical / education / arts | Low |
| S_BL_OCC_3D_EN051 | Ever/Never Science/Tech Administration Persnl | Low | Professional / technical / medical / education / arts | Low |
| S_BL_OCC_3D_EN052 | Ever/Never Science/Tech Supporting Persnl | Low | Professional / technical / medical / education / arts | Low |
| S_BL_OCC_3D_EN061 | Ever/Never Aircraft Pilots and Navigators | Low | Professional / technical / medical / education / arts | Low |
| S_BL_OCC_3D_EN062 | Ever/Never Aircraft Mechanics | Low | Professional / technical / medical / education / arts | Low |
| S_BL_OCC_3D_EN063 | Ever/Never Ship Commanders and Navigators | Low | Professional / technical / medical / education / arts | Low |
| S_BL_OCC_3D_EN064 | Ever/Never Ship's Turbine Technology Persnl | Low | Professional / technical / medical / education / arts | Low |
| S_BL_OCC_3D_EN069 | Ever/Never Tech Persnl in Aircraft & Ships | Low | Professional / technical / medical / education / arts | Low |
| S_BL_OCC_3D_EN071 | Ever/Never Practitioners Chinese Trad Med | Low | Professional / technical / medical / education / arts | Low |
| S_BL_OCC_3D_EN072 | Ever/Never Practitioners of Western Med | Low | Professional / technical / medical / education / arts | Low |
| S_BL_OCC_3D_EN073 | Ever/Never Practit Chinese Trad Med & West Med | Low | Professional / technical / medical / education / arts | Low |
| S_BL_OCC_3D_EN074 | Ever/Never Pharmacists | Low | Professional / technical / medical / education / arts | Low |
| S_BL_OCC_3D_EN075 | Ever/Never Nurses | Low | Professional / technical / medical / education / arts | Low |
| S_BL_OCC_3D_EN076 | Ever/Never Technical Experts | Low | Professional / technical / medical / education / arts | Low |
| S_BL_OCC_3D_EN078 | Ever/Never Urban Health Technology Persnl | Low | Professional / technical / medical / education / arts | Low |
| S_BL_OCC_3D_EN089 | Ever/Never Other Med/Health Tech Persnl | Low | Professional / technical / medical / education / arts | Low |
| S_BL_OCC_3D_EN091 | Ever/Never Economy Planners | Low | Professional / technical / medical / education / arts | Low |
| S_BL_OCC_3D_EN092 | Ever/Never Statisticians | Low | Professional / technical / medical / education / arts | Low |
| S_BL_OCC_3D_EN093 | Ever/Never Accounting Personnel | Low | Professional / technical / medical / education / arts | Low |
| S_BL_OCC_3D_EN094 | Ever/Never Personnel Workers | Low | Professional / technical / medical / education / arts | Low |
| S_BL_OCC_3D_EN095 | Ever/Never Dispatchers | Low | Professional / technical / medical / education / arts | Low |
| S_BL_OCC_3D_EN096 | Ever/Never Tax Collectrs/Admin Indust/Commerce | Low | Professional / technical / medical / education / arts | Low |
| S_BL_OCC_3D_EN097 | Ever/Never Finance Workers | Low | Professional / technical / medical / education / arts | Low |
| S_BL_OCC_3D_EN098 | Ever/Never Customs Inspectors | Low | Professional / technical / medical / education / arts | Low |
| S_BL_OCC_3D_EN099 | Ever/Never Other Persnl Economics/Finance | Low | Professional / technical / medical / education / arts | Low |
| S_BL_OCC_3D_EN101 | Ever/Never Judges | Low | Professional / technical / medical / education / arts | Low |
| S_BL_OCC_3D_EN102 | Ever/Never Prosecutors | Low | Professional / technical / medical / education / arts | Low |
| S_BL_OCC_3D_EN103 | Ever/Never Lawyers | Low | Professional / technical / medical / education / arts | Low |
| S_BL_OCC_3D_EN109 | Ever/Never Other Personnel Engaged in Law | Low | Professional / technical / medical / education / arts | Low |
| S_BL_OCC_3D_EN111 | Ever/Never University-level Teachers | Low | Professional / technical / medical / education / arts | Low |
| S_BL_OCC_3D_EN112 | Ever/Never Middle-level Teachers | Low | Professional / technical / medical / education / arts | Low |
| S_BL_OCC_3D_EN113 | Ever/Never Elementary-level Teachers | Low | Professional / technical / medical / education / arts | Low |
| S_BL_OCC_3D_EN114 | Ever/Never Nursery School Teachers | Low | Professional / technical / medical / education / arts | Low |
| S_BL_OCC_3D_EN115 | Ever/Never Teachers of Blind, Deaf &/or Mute | Low | Professional / technical / medical / education / arts | Low |
| S_BL_OCC_3D_EN116 | Ever/Never Teachers' Aids | Low | Professional / technical / medical / education / arts | Low |
| S_BL_OCC_3D_EN119 | Ever/Never Other Teachers | Low | Professional / technical / medical / education / arts | Low |
| S_BL_OCC_3D_EN121 | Ever/Never Lit/Arts Creation/Critics/Reviews | Low | Professional / technical / medical / education / arts | Low |
| S_BL_OCC_3D_EN122 | Ever/Never Editors/Directrs in Perform Arts | Low | Professional / technical / medical / education / arts | Low |
| S_BL_OCC_3D_EN123 | Ever/Never Performers | Low | Professional / technical / medical / education / arts | Low |
| S_BL_OCC_3D_EN124 | Ever/Never Musical Instruments Performers | Low | Professional / technical / medical / education / arts | Low |
| S_BL_OCC_3D_EN125 | Ever/Never Photographers | Low | Professional / technical / medical / education / arts | Low |
| S_BL_OCC_3D_EN126 | Ever/Never Movies/TV Cameramen | Low | Professional / technical / medical / education / arts | Low |
| S_BL_OCC_3D_EN127 | Ever/Never Fine Artists | Low | Professional / technical / medical / education / arts | Low |
| S_BL_OCC_3D_EN128 | Ever/Never Stage Workers | Low | Professional / technical / medical / education / arts | Low |
| S_BL_OCC_3D_EN129 | Ever/Never Sports Coaches and Umpires | Low | Professional / technical / medical / education / arts | Low |
| S_BL_OCC_3D_EN130 | Ever/Never Athletes | Low | Professional / technical / medical / education / arts | Low |
| S_BL_OCC_3D_EN139 | Ever/Never Other Literary/Arts/Sports Persnl | Low | Professional / technical / medical / education / arts | Low |
| S_BL_OCC_3D_EN141 | Ever/Never Reporters | Low | Professional / technical / medical / education / arts | Low |
| S_BL_OCC_3D_EN142 | Ever/Never Editors | Low | Professional / technical / medical / education / arts | Low |
| S_BL_OCC_3D_EN143 | Ever/Never Broadcasters | Low | Professional / technical / medical / education / arts | Low |
| S_BL_OCC_3D_EN144 | Ever/Never Translators & Interpreters | Low | Professional / technical / medical / education / arts | Low |
| S_BL_OCC_3D_EN145 | Ever/Never Librarians & Library Workers | Low | Professional / technical / medical / education / arts | Low |
| S_BL_OCC_3D_EN146 | Ever/Never Files/Archives Management Workrs | Low | Professional / technical / medical / education / arts | Low |
| S_BL_OCC_3D_EN147 | Ever/Never Archeol/Cultural Relics Workrs | Low | Professional / technical / medical / education / arts | Low |
| S_BL_OCC_3D_EN148 | Ever/Never Exhibition Tour Guides | Low | Professional / technical / medical / education / arts | Low |
| S_BL_OCC_3D_EN149 | Ever/Never Other Cultural Workers | Low | Professional / technical / medical / education / arts | Low |
| S_BL_OCC_3D_EN150 | Ever/Never Professional Religious Workers | Low | Professional / technical / medical / education / arts | Low |
| S_BL_OCC_3D_EN211 | Ever/Never In Charge Central Gov Bodies | Low | Administrative / managerial | Low |
| S_BL_OCC_3D_EN212 | Ever/Never In Charge Prov-Level State Bodies | Low | Administrative / managerial | Low |
| S_BL_OCC_3D_EN213 | Ever/Never In Charge Reg-/City-/Coun-Level St | Low | Administrative / managerial | Low |
| S_BL_OCC_3D_EN221 | Ever/Never In Charge Chinese Communist Party | Low | Administrative / managerial | Low |
| S_BL_OCC_3D_EN222 | Ever/Never Charge Chinese Communist Youth Lgue | Low | Administrative / managerial | Low |
| S_BL_OCC_3D_EN223 | Ever/Never In Charge Chinese Democracy Party | Low | Administrative / managerial | Low |
| S_BL_OCC_3D_EN231 | Ever/Never In Charge Urban Neighborhoods | Low | Administrative / managerial | Low |
| S_BL_OCC_3D_EN232 | Ever/Never In Charge of Res Committees | Low | Administrative / managerial | Low |
| S_BL_OCC_3D_EN233 | Ever/Never In Charge of Rural Communes | Low | Administrative / managerial | Low |
| S_BL_OCC_3D_EN241 | Ever/Never In Charge Enterprise/Instit Units | Low | Administrative / managerial | Low |
| S_BL_OCC_3D_EN242 | Ever/Never In Charge Wrk Orgs/Enterprise Units | Low | Administrative / managerial | Low |
| S_BL_OCC_3D_EN311 | Ever/Never Office Workers in Administration | Low | Clerical / security / postal / telecom | Low |
| S_BL_OCC_3D_EN312 | Ever/Never Secretaries | Low | Clerical / security / postal / telecom | Low |
| S_BL_OCC_3D_EN313 | Ever/Never Mail Clerks & Communic Workers | Low | Clerical / security / postal / telecom | Low |
| S_BL_OCC_3D_EN314 | Ever/Never Typists, Mimeographers | Low | Clerical / security / postal / telecom | Low |
| S_BL_OCC_3D_EN319 | Ever/Never Other Admin Office Workers | Low | Clerical / security / postal / telecom | Low |
| S_BL_OCC_3D_EN320 | Ever/Never Armymen | Low | Clerical / security / postal / telecom | Low |
| S_BL_OCC_3D_EN321 | Ever/Never Political Workers | Low | Clerical / security / postal / telecom | Low |
| S_BL_OCC_3D_EN322 | Ever/Never Persnl [cadre] Management Workrs | Low | Clerical / security / postal / telecom | Low |
| S_BL_OCC_3D_EN323 | Ever/Never Policemen/Pub Security Workers | Low | Clerical / security / postal / telecom | Low |
| S_BL_OCC_3D_EN324 | Ever/Never Firemen | Low | Clerical / security / postal / telecom | Low |
| S_BL_OCC_3D_EN329 | Ever/Never Other Polit & Security Workers | Low | Clerical / security / postal / telecom | Low |
| S_BL_OCC_3D_EN331 | Ever/Never Postal Service Workers | Low | Clerical / security / postal / telecom | Low |
| S_BL_OCC_3D_EN332 | Ever/Never Telecommunication Service Workrs | Low | Clerical / security / postal / telecom | Low |
| S_BL_OCC_3D_EN339 | Ever/Never Other Postal/Telecomm Workers | Low | Clerical / security / postal / telecom | Low |
| S_BL_OCC_3D_EN391 | Ever/Never Non-Technical Managing Personnel | Low | Clerical / security / postal / telecom | Low |
| S_BL_OCC_3D_EN399 | Ever/Never Other Affairs Handler Workers | Low | Clerical / security / postal / telecom | Low |
| S_BL_OCC_3D_EN411 | Ever/Never Business Operators, Sales Persons | Low | Commerce / sales / purchasing | Low |
| S_BL_OCC_3D_EN419 | Ever/Never Other Persons Engaged in Sales | Low | Commerce / sales / purchasing | Low |
| S_BL_OCC_3D_EN421 | Ever/Never Purchasing Agents | Low | Commerce / sales / purchasing | Low |
| S_BL_OCC_3D_EN422 | Ever/Never Persons in Supplying/Marketing | Low | Commerce / sales / purchasing | Low |
| S_BL_OCC_3D_EN432 | Ever/Never Buyers on CONSIGNMENTS | Low | Commerce / sales / purchasing | Low |
| S_BL_OCC_3D_EN499 | Ever/Never Other Persons in Business | Low | Commerce / sales / purchasing | Low |
| S_BL_OCC_3D_EN511 | Ever/Never Service Persnl in Restaurants | Low | General service work | Low |
| S_BL_OCC_3D_EN512 | Ever/Never Service Hotels/Guest Houses/etc | Low | General service work | Low |
| S_BL_OCC_3D_EN513 | Ever/Never Service Theaters/Pub Displays | Low | General service work | Low |
| S_BL_OCC_3D_EN514 | Ever/Never Service Persnl in Bath Houses | Low | General service work | Low |
| S_BL_OCC_3D_EN515 | Ever/Never Service Vehicles/Ships/Airplanes | Low | General service work | Low |
| S_BL_OCC_3D_EN516 | Ever/Never Ticket Sellers | Low | General service work | Low |
| S_BL_OCC_3D_EN517 | Ever/Never Nursery Child Care Workers | Low | General service work | Low |
| S_BL_OCC_3D_EN518 | Ever/Never Nannies | Low | General service work | Low |
| S_BL_OCC_3D_EN519 | Ever/Never Barbers & Hairdressers | Low | General service work | Low |
| S_BL_OCC_3D_EN520 | Ever/Never Laundering/Dyeing/Repair Garments | Low | General service work | Low |
| S_BL_OCC_3D_EN521 | Ever/Never Suppliers of Fuels | Low | General service work | Low |
| S_BL_OCC_3D_EN523 | Ever/Never Odds-and-Ends Workers | Low | General service work | Low |
| S_BL_OCC_3D_EN529 | Ever/Never Other Service Personnel | Low | General service work | Low |
| S_BL_OCC_3D_EN530 | Ever/Never Chefs and Cooks | Low | General service work | Low |
| S_BL_OCC_3D_EN540 | Ever/Never Tour Guides | Low | General service work | Low |
| S_BL_OCC_3D_EN551 | Ever/Never Repairers Mech/Elec Consumer Prods | Low | General service work | Low |
| S_BL_OCC_3D_EN559 | Ever/Never Other Repairers Everyday Articles | Low | General service work | Low |
| S_BL_OCC_3D_EN599 | Ever/Never Other Service Personnel | Low | General service work | Low |
| S_BL_OCC_3D_EN623 | Ever/Never Forest Protection Personnel | Low | Forestry | Medium |
| S_BL_OCC_3D_EN650 | Ever/Never Hunters | Low | Hunting | Low |
| S_BL_OCC_3D_EN711 | Ever/Never Mining, Quarrying | Low | Mining / metals / chemicals / rubber / plastics | Low |
| S_BL_OCC_3D_EN712 | Ever/Never Ores and Rocks Processing | Low | Mining / metals / chemicals / rubber / plastics | Low |
| S_BL_OCC_3D_EN713 | Ever/Never Extraction of Oil, Natural Gas | Low | Mining / metals / chemicals / rubber / plastics | Low |
| S_BL_OCC_3D_EN714 | Ever/Never Geological Exploration | Low | Mining / metals / chemicals / rubber / plastics | Low |
| S_BL_OCC_3D_EN715 | Ever/Never Salt-Mining | Low | Mining / metals / chemicals / rubber / plastics | Low |
| S_BL_OCC_3D_EN719 | Ever/Never Other Mining/Quarrying/Exploration | Low | Mining / metals / chemicals / rubber / plastics | Low |
| S_BL_OCC_3D_EN721 | Ever/Never Smelters | Low | Mining / metals / chemicals / rubber / plastics | Low |
| S_BL_OCC_3D_EN722 | Ever/Never Rolling Mill/Machinery Operators | Low | Mining / metals / chemicals / rubber / plastics | Low |
| S_BL_OCC_3D_EN723 | Ever/Never Foundry Workers | Low | Mining / metals / chemicals / rubber / plastics | Low |
| S_BL_OCC_3D_EN724 | Ever/Never Heat Treatment of Metals | Low | Mining / metals / chemicals / rubber / plastics | Low |
| S_BL_OCC_3D_EN725 | Ever/Never Drawing & Extruding of Metals | Low | Mining / metals / chemicals / rubber / plastics | Low |
| S_BL_OCC_3D_EN726 | Ever/Never Surface Treatment of Metals | Low | Mining / metals / chemicals / rubber / plastics | Low |
| S_BL_OCC_3D_EN729 | Ever/Never Other Ore Smelting/Metal Process | Low | Mining / metals / chemicals / rubber / plastics | Low |
| S_BL_OCC_3D_EN731 | Ever/Never Basic Chem Raw Materials Productn | Low | Mining / metals / chemicals / rubber / plastics | Low |
| S_BL_OCC_3D_EN732 | Ever/Never Chemical Fertilizers Production | Low | Mining / metals / chemicals / rubber / plastics | Low |
| S_BL_OCC_3D_EN733 | Ever/Never Farm Chemicals Production Workers | Low | Mining / metals / chemicals / rubber / plastics | Low |
| S_BL_OCC_3D_EN734 | Ever/Never Everyday Use Chemicals Production | Low | Mining / metals / chemicals / rubber / plastics | Low |
| S_BL_OCC_3D_EN735 | Ever/Never Chemical Fibers Production Workers | Low | Mining / metals / chemicals / rubber / plastics | Low |
| S_BL_OCC_3D_EN736 | Ever/Never Oil Refinery Workers | Low | Mining / metals / chemicals / rubber / plastics | Low |
| S_BL_OCC_3D_EN737 | Ever/Never Coking Workers | Low | Mining / metals / chemicals / rubber / plastics | Low |
| S_BL_OCC_3D_EN738 | Ever/Never Pharmaceutical Workers | Low | Mining / metals / chemicals / rubber / plastics | Low |
| S_BL_OCC_3D_EN739 | Ever/Never Other Chemicals Workers | Low | Mining / metals / chemicals / rubber / plastics | Low |
| S_BL_OCC_3D_EN741 | Ever/Never Rubber Process Machine Operators | Low | Mining / metals / chemicals / rubber / plastics | Low |
| S_BL_OCC_3D_EN742 | Ever/Never Plastic Process Machine Operators | Low | Mining / metals / chemicals / rubber / plastics | Low |
| S_BL_OCC_3D_EN743 | Ever/Never Tire Production/Vulcanization | Low | Mining / metals / chemicals / rubber / plastics | Low |
| S_BL_OCC_3D_EN749 | Ever/Never Other Rubber/Plastic Production | Low | Mining / metals / chemicals / rubber / plastics | Low |
| S_BL_OCC_3D_EN803 | Ever/Never Wood Furniture Workers | Low | Wood / paper processing | Medium |
| S_BL_OCC_3D_EN804 | Ever/Never Construction Carpenters | Low | Wood / paper processing | Medium |
| S_BL_OCC_3D_EN821 | Ever/Never Character Selection & Typesetting | Low | Printing / plate making | Low |
| S_BL_OCC_3D_EN822 | Ever/Never Plate Makers | Low | Printing / plate making | Low |
| S_BL_OCC_3D_EN823 | Ever/Never Printing Workers | Low | Printing / plate making | Low |
| S_BL_OCC_3D_EN824 | Ever/Never Binders | Low | Printing / plate making | Low |
| S_BL_OCC_3D_EN825 | Ever/Never Dark Room Photography Workers | Low | Printing / plate making | Low |
| S_BL_OCC_3D_EN829 | Ever/Never Other Printers & Related Workers | Low | Printing / plate making | Low |
| S_BL_OCC_3D_EN831 | Ever/Never Stonecutters | Low | Stone / glass / ceramics | Low |
| S_BL_OCC_3D_EN832 | Ever/Never Stone Craftsmen | Low | Stone / glass / ceramics | Low |
| S_BL_OCC_3D_EN839 | Ever/Never Other Stone Workers | Low | Stone / glass / ceramics | Low |
| S_BL_OCC_3D_EN841 | Ever/Never Forgers & Forging Press Operators | Low | Machinery / electrical / electronic installation & maintenance | Low |
| S_BL_OCC_3D_EN842 | Ever/Never Fitters | Low | Machinery / electrical / electronic installation & maintenance | Low |
| S_BL_OCC_3D_EN843 | Ever/Never Machine Tool Installers | Low | Machinery / electrical / electronic installation & maintenance | Low |
| S_BL_OCC_3D_EN844 | Ever/Never Metal-cut Mach Tools/Blade Sharpen | Low | Machinery / electrical / electronic installation & maintenance | Low |
| S_BL_OCC_3D_EN849 | Ever/Never Other Forge/Tool/Mach Tool Install | Low | Machinery / electrical / electronic installation & maintenance | Low |
| S_BL_OCC_3D_EN851 | Ever/Never Machinery Installation & Assembly | Low | Machinery / electrical / electronic installation & maintenance | Low |
| S_BL_OCC_3D_EN852 | Ever/Never Machinery Maintenance Workers | Low | Machinery / electrical / electronic installation & maintenance | Low |
| S_BL_OCC_3D_EN853 | Ever/Never Clocks & Watches Instrument Prod | Low | Machinery / electrical / electronic installation & maintenance | Low |
| S_BL_OCC_3D_EN859 | Ever/Never Other Instal/Assembly Mach Product | Low | Machinery / electrical / electronic installation & maintenance | Low |
| S_BL_OCC_3D_EN861 | Ever/Never Install/Maint Electric Equip | Low | Machinery / electrical / electronic installation & maintenance | Low |
| S_BL_OCC_3D_EN862 | Ever/Never Install/Maint Electronic Equip | Low | Machinery / electrical / electronic installation & maintenance | Low |
| S_BL_OCC_3D_EN863 | Ever/Never Assembly Electric/Electron Equip | Low | Machinery / electrical / electronic installation & maintenance | Low |
| S_BL_OCC_3D_EN864 | Ever/Never Electricians | Low | Machinery / electrical / electronic installation & maintenance | Low |
| S_BL_OCC_3D_EN865 | Ever/Never Instal/Maint Phone/Telegraph Equip | Low | Machinery / electrical / electronic installation & maintenance | Low |
| S_BL_OCC_3D_EN869 | Ever/Never Oth Elect Equip Instal/Maint | Low | Machinery / electrical / electronic installation & maintenance | Low |
| S_BL_OCC_3D_EN871 | Ever/Never Transmission Equipment Operators | Low | Equipment operation / media / tape production | Low |
| S_BL_OCC_3D_EN872 | Ever/Never Sound Recording Equipment Operators | Low | Equipment operation / media / tape production | Low |
| S_BL_OCC_3D_EN873 | Ever/Never Motion Picture Projector Operators | Low | Equipment operation / media / tape production | Low |
| S_BL_OCC_3D_EN874 | Ever/Never Tape Making Workers | Low | Equipment operation / media / tape production | Low |
| S_BL_OCC_3D_EN881 | Ever/Never Workers in Ductworks and Pipelines | Low | Metal fabrication / welding / ductwork | Low |
| S_BL_OCC_3D_EN882 | Ever/Never Welders | Low | Metal fabrication / welding / ductwork | Low |
| S_BL_OCC_3D_EN883 | Ever/Never Cold Working Workers | Low | Metal fabrication / welding / ductwork | Low |
| S_BL_OCC_3D_EN884 | Ever/Never Makers/Installers Metal Components | Low | Metal fabrication / welding / ductwork | Low |
| S_BL_OCC_3D_EN891 | Ever/Never Glass Workers | Low | Glass / pottery / enamel | Low |
| S_BL_OCC_3D_EN892 | Ever/Never Pottery and Porcelain Workers | Low | Glass / pottery / enamel | Low |
| S_BL_OCC_3D_EN893 | Ever/Never Enamel Workers | Low | Glass / pottery / enamel | Low |
| S_BL_OCC_3D_EN901 | Ever/Never Painters | Low | Painting / non-metal mineral / other production | Low |
| S_BL_OCC_3D_EN909 | Ever/Never Other Painters | Low | Painting / non-metal mineral / other production | Low |
| S_BL_OCC_3D_EN911 | Ever/Never Production Cement & Cement Products | Low | Painting / non-metal mineral / other production | Low |
| S_BL_OCC_3D_EN912 | Ever/Never Prod Asbestos & Asbestos Products | Low | Painting / non-metal mineral / other production | Low |
| S_BL_OCC_3D_EN913 | Ever/Never Production Bricks/Tiles/Lime | Low | Painting / non-metal mineral / other production | Low |
| S_BL_OCC_3D_EN914 | Ever/Never Production Refractory Materials | Low | Painting / non-metal mineral / other production | Low |
| S_BL_OCC_3D_EN915 | Ever/Never Prod Other Non-Metal Mineral Prods | Low | Painting / non-metal mineral / other production | Low |
| S_BL_OCC_3D_EN916 | Ever/Never Prod Cultural/Educ/Sports Articles | Low | Painting / non-metal mineral / other production | Low |
| S_BL_OCC_3D_EN917 | Ever/Never Jewelers | Low | Painting / non-metal mineral / other production | Low |
| S_BL_OCC_3D_EN918 | Ever/Never Engravers of Artistic Works | Low | Painting / non-metal mineral / other production | Low |
| S_BL_OCC_3D_EN919 | Ever/Never Other Production & Related Workers | Low | Painting / non-metal mineral / other production | Low |
| S_BL_OCC_3D_EN921 | Ever/Never Tile Workers, Plasterers | Low | Construction | Low |
| S_BL_OCC_3D_EN922 | Ever/Never Concrete Workers | Low | Construction | Low |
| S_BL_OCC_3D_EN923 | Ever/Never Steel Benders | Low | Construction | Low |
| S_BL_OCC_3D_EN924 | Ever/Never Scaffolders | Low | Construction | Low |
| S_BL_OCC_3D_EN925 | Ever/Never Laborers Engaged in Heavy Work | Low | Construction | Low |
| S_BL_OCC_3D_EN929 | Ever/Never Other Construction Workers | Low | Construction | Low |
| S_BL_OCC_3D_EN931 | Ever/Never Electricity Generation Equipment | Low | Power generation / boiler operations | Low |
| S_BL_OCC_3D_EN932 | Ever/Never Around-the-Clock Power Networks | Low | Power generation / boiler operations | Low |
| S_BL_OCC_3D_EN933 | Ever/Never Boiler Workers | Low | Power generation / boiler operations | Low |
| S_BL_OCC_3D_EN939 | Ever/Never Other Power Generation Equipment | Low | Power generation / boiler operations | Low |
| S_BL_OCC_3D_EN941 | Ever/Never Loading/Unloading Workers & Movers | Low | Loading / transport / shipping | Low |
| S_BL_OCC_3D_EN942 | Ever/Never Crane Operators | Low | Loading / transport / shipping | Low |
| S_BL_OCC_3D_EN943 | Ever/Never Earth-moving Mach & Related Equip | Low | Loading / transport / shipping | Low |
| S_BL_OCC_3D_EN949 | Ever/Never Other Loading/Unloading Workers | Low | Loading / transport / shipping | Low |
| S_BL_OCC_3D_EN951 | Ever/Never Ship Crew | Low | Loading / transport / shipping | Low |
| S_BL_OCC_3D_EN952 | Ever/Never Locomotive Drivers & Stokers | Low | Loading / transport / shipping | Low |
| S_BL_OCC_3D_EN953 | Ever/Never Railroad Transportation Workers | Low | Loading / transport / shipping | Low |
| S_BL_OCC_3D_EN954 | Ever/Never Motor Vehicle Drivers | Low | Loading / transport / shipping | Low |
| S_BL_OCC_3D_EN955 | Ever/Never Power-driven Vehicle Drivers | Low | Loading / transport / shipping | Low |
| S_BL_OCC_3D_EN957 | Ever/Never Man-powered Vehicle Workers | Low | Loading / transport / shipping | Low |
| S_BL_OCC_3D_EN959 | Ever/Never Other Equipment Used in Transport | Low | Loading / transport / shipping | Low |
| S_BL_OCC_3D_EN961 | Ever/Never Examiners/Inspectors | Low | Inspection / testing / laboratory analysis | Low |
| S_BL_OCC_3D_EN962 | Ever/Never Measurers | Low | Inspection / testing / laboratory analysis | Low |
| S_BL_OCC_3D_EN966 | Ever/Never Other Examiners | Low | Inspection / testing / laboratory analysis | Low |
| S_BL_OCC_3D_EN967 | Ever/Never Chemical Testers | Low | Inspection / testing / laboratory analysis | Low |
| S_BL_OCC_3D_EN991 | Ever/Never Workers in Underwater Operations | Low | Miscellaneous / warehouse / underwater | Low |
| S_BL_OCC_3D_EN992 | Ever/Never Mapping Workers | Low | Miscellaneous / warehouse / underwater | Low |
| S_BL_OCC_3D_EN993 | Ever/Never Workers in Packaging/Baling | Low | Miscellaneous / warehouse / underwater | Low |
| S_BL_OCC_3D_EN994 | Ever/Never Warehouse Workers | Low |  |  |
| S_BL_OCC_3D_EN997 | Ever/Never Other Production/Transport Workers | Low |  |  |
| S_BL_OCC_3D_EN998 | Ever/Never Miscellaneous other workers | Low |  |  |
| S_BL_OCC_3D_EN019 | Ever/Never Researchers in Science | Medium | Professional / technical / medical / education / arts | Low |
| S_BL_OCC_3D_EN021 | Ever/Never Researchers in Agriculture | Medium | Professional / technical / medical / education / arts | Low |
| S_BL_OCC_3D_EN022 | Ever/Never Researchers in Medicine | Medium | Professional / technical / medical / education / arts | Low |
| S_BL_OCC_3D_EN042 | Ever/Never Light Indust & Textile Tech Persnl | Medium | Professional / technical / medical / education / arts | Low |
| S_BL_OCC_3D_EN043 | Ever/Never Agriculture Technology Personnel | Medium | Professional / technical / medical / education / arts | Low |
| S_BL_OCC_3D_EN044 | Ever/Never Forestry Technology Personnel | Medium | Professional / technical / medical / education / arts | Low |
| S_BL_OCC_3D_EN049 | Ever/Never Other Tech Persnl Eng/Agric/Forstry | Medium | Professional / technical / medical / education / arts | Low |
| S_BL_OCC_3D_EN077 | Ever/Never Persnl Hygiene/Anti-Epidemic Work | Medium | Professional / technical / medical / education / arts | Low |
| S_BL_OCC_3D_EN431 | Ever/Never Buyers of Farm/Sideline Products | Medium | Commerce / sales / purchasing | Low |
| S_BL_OCC_3D_EN524 | Ever/Never Funeral and Burial Workers | Medium | General service work | Low |
| S_BL_OCC_3D_EN621 | Ever/Never Forest Raising/Planting Persnl | Medium | Forestry | Medium |
| S_BL_OCC_3D_EN622 | Ever/Never Tree Felling Personnel | Medium | Forestry | Medium |
| S_BL_OCC_3D_EN624 | Ever/Never Water Transport of Timber | Medium | Forestry | Medium |
| S_BL_OCC_3D_EN629 | Ever/Never Other Forestry Workers | Medium | Forestry | Medium |
| S_BL_OCC_3D_EN641 | Ever/Never Aquatic Plants/Animals Raisers | Medium | Fishery / aquatic products | Medium |
| S_BL_OCC_3D_EN642 | Ever/Never Fishing/Capturing Aquatic Animals | Medium | Fishery / aquatic products | Medium |
| S_BL_OCC_3D_EN643 | Ever/Never Collect/Harvest Natural Aqua Prod | Medium | Fishery / aquatic products | Medium |
| S_BL_OCC_3D_EN644 | Ever/Never Operators Motorized Fishing Boats | Medium | Fishery / aquatic products | Medium |
| S_BL_OCC_3D_EN649 | Ever/Never Other Fish/Captre/Harvst Aqua Prods | Medium | Fishery / aquatic products | Medium |
| S_BL_OCC_3D_EN762 | Ever/Never Production finished leather goods | Medium | Leather / fur / feather processing | Medium |
| S_BL_OCC_3D_EN763 | Ever/Never Production finished fur goods | Medium | Leather / fur / feather processing | Medium |
| S_BL_OCC_3D_EN769 | Ever/Never Other Leathers/Furs/Finished Prod | Medium | Leather / fur / feather processing | Medium |
| S_BL_OCC_3D_EN771 | Ever/Never Pattern Makers, Tailoring Workers | Medium | Tailoring / sewing | Medium |
| S_BL_OCC_3D_EN772 | Ever/Never Sewing Workers | Medium | Tailoring / sewing | Medium |
| S_BL_OCC_3D_EN773 | Ever/Never Shoemakers, Hat-Makers | Medium | Tailoring / sewing | Medium |
| S_BL_OCC_3D_EN774 | Ever/Never Embroidery Workers | Medium | Tailoring / sewing | Medium |
| S_BL_OCC_3D_EN779 | Ever/Never Other Tailoring and Sewing Workers | Medium | Tailoring / sewing | Medium |
| S_BL_OCC_3D_EN782 | Ever/Never Sugar Production Workers | Medium | Raw food / grain / animal-food processing | High |
| S_BL_OCC_3D_EN784 | Ever/Never Pickling/Canning/Preserv Food | Medium | Raw food / grain / animal-food processing | High |
| S_BL_OCC_3D_EN785 | Ever/Never Processing Milk Products | Medium | Raw food / grain / animal-food processing | High |
| S_BL_OCC_3D_EN786 | Ever/Never Bakers, Confectioners | Medium | Other food / beverage / tobacco processing | Medium |
| S_BL_OCC_3D_EN788 | Ever/Never Wine-Makers/Brewers/Beverage Prod | Medium | Other food / beverage / tobacco processing | Medium |
| S_BL_OCC_3D_EN789 | Ever/Never Other Food/Beverage Production | Medium | Other food / beverage / tobacco processing | Medium |
| S_BL_OCC_3D_EN802 | Ever/Never Process Timber into Construct Prod | Medium | Wood / paper processing | Medium |
| S_BL_OCC_3D_EN809 | Ever/Never Other Timber/Finished Prods | Medium | Wood / paper processing | Medium |
| S_BL_OCC_3D_EN811 | Ever/Never Paper-making Workers | Medium | Wood / paper processing | Medium |
| S_BL_OCC_3D_EN812 | Ever/Never Paper Products Workers | Medium | Wood / paper processing | Medium |
| S_BL_OCC_3D_EN819 | Ever/Never Other Paper-making & Paper Prod | Medium | Wood / paper processing | Medium |
| S_BL_OCC_3D_EN956 | Ever/Never Drivers of Animal-Powered Vehicles | Medium | Loading / transport / shipping | Low |
| S_BL_OCC_3D_EN963 | Ever/Never Testers/Experimenters | Medium | Inspection / testing / laboratory analysis | Low |
| S_BL_OCC_3D_EN964 | Ever/Never Lab Technicians/Analyzers | Medium | Inspection / testing / laboratory analysis | Low |
| S_BL_OCC_3D_EN965 | Ever/Never Chemical Examiners | Medium | Inspection / testing / laboratory analysis | Low |

